# Drug-coated balloons for dysfunctional hemodialysis venous access: A patient-level meta-analysis of randomized controlled trials

**DOI:** 10.1101/2020.12.10.20240069

**Authors:** Fong Khi Yung, Joseph J Zhao, Eelin Tan, Nicholas Syn, Rehena Sultana, Kun Da Zhuang, Jasmine Chua, Er Ming, Ankur Patel, Farah Gillan Irani, Tay Kiang Hiong, Tan Bien Soo, Too Chow Wei

## Abstract

**Purpose:** To perform an individual patient data-level meta-analysis of randomized controlled trials comparing drug-coated balloon angioplasty (DCB) against conventional percutaneous transluminal angioplasty (PTA) in the treatment of dysfunctional hemodialysis venous access.

**Methods:** A search was conducted from inception till 13^th^ November 2020. Kaplan-Meier curves comparing DCB to PTA by target lesion primary patency (TLPP) and access circuit primary patency (ACPP) were graphically reconstructed to retrieve patient-level data. One-stage meta-analyses with Cox-models with random-effects gramma-frailties were conducted to determine hazard ratios (HRs). Dynamic restricted mean survival times (RMST) were conducted in view of violation of the proportional hazards assumption. Conventional two-stage meta-analyses and network meta-analyses under random-effects Frequentist models were conducted to determine overall and comparative outcomes of paclitaxel concentrations utilised. Where outliers were consistently detected through outlier and influence analyses, sensitivity analyses excluding those studies were conducted.

**Results:** Among 10 RCTs (1,207 patients), HRs across all models favoured DCB (one-stage shared-frailty HR=0.62, 95%-CI: 0.53–0.73, P<0.001; two-stage random-effects HR=0.60, 95%-CI: 0.42–0.86, P=0.018, *I*^*2*^=65%) for TLPP. Evidence of time-varying effects (P=0.005) was found. TLPP RMST was +3.47 months (25.0%) longer in DCB-treated patients compared to PTA (P=0.001) at 3-years. TLPP at 6-months, 1-year and 2-years was 75.3% vs 58.0%, 51.1% vs 37.1% and 31.3% vs 26.0% for DCB and PTA respectively. P-Scores within the Frequentist network meta-analysis suggest that higher concentrations of paclitaxel were associated with better TLPP and ACPP. Among 6 RCTs (854 patients), the one-stage model favoured DCB (shared-frailty HR=0.72, 95%-CI: 0.60–0.87, P<0.001) for ACPP. Conversely, the two-stage random-effects model demonstrated no significant difference (HR=0.76, 95%-CI: 0.35–1.67, P=0.414, *I*^*2*^=81%). Sensitivity analysis excluding outliers significantly favoured DCB (HR=0.61, 95%-CI: 0.41–0.91, P=0.027, *I*^*2*^=62%).

**Conclusion:** Overall evidence suggests that DCB is favoured over PTA in TLPP and ACPP. The increased efficacy of higher concentrations of paclitaxel may warrant further investigation.

## Introduction

Venous stenosis is the commonest cause of dysfunction in arteriovenous fistulas (AVFs) and arteriovenous grafts (AVGs) used for hemodialysis [1]. The Kidney Disease Outcomes Quality Initiative (KDOQI) recommends percutaneous transluminal angioplasty (PTA) as first-line treatment to restore vessel patency [2]. While PTA procedural success is generally high, microscopic damage to the vessel wall upon inflation may cause reactive intimal hyperplasia [3-5], inducing restenosis and poor long-term patency [6].

Presently, drug-coated balloon (DCB) angioplasty has become an increasing subject of discussion as an adjunct to PTA. DCBs are commonly coated with paclitaxel, which prevents neointimal hyperplasia in blood vessels [7-9].

Paclitaxel DCB has been demonstrated to be superior to PTA in the contexts of femoropopliteal artery disease [10] and coronary instent restenosis [11]. However, findings are mixed in the setting of hemodialysis access – some meta-analyses significantly favoured DCB over PTA [12, 13], while others reported no significant benefit [14, 15]. Not long ago, two new randomized controlled trials (RCTs) on this subject were published [16, 17]; this article aims to re-evaluate the field with these new inclusions. To our knowledge, this is first meta-analysis which includes these 2 new RCTs.

In lieu of this area’s uncertain and developing nature, more precise methods are needed to quantify the comparison between DCB and PTA. Thus, we utilised graphical reconstructive algorithms to attain survival information of individual patients [18, 19] from Kaplan-Meier curves. Individual patient data (IPD) is recognised as the gold standard approach for evidence synthesis [20, 21]. The source of IPD for this study was restricted to RCTs only, as baseline characteristic differences between patients and differing indications for treatment render non-randomized studies difficult to interpret definitively.

Leveraging the IPD of 11 RCTs, this meta-analysis aims to compare the relative efficacy of DCB to PTA in maintaining target lesion primary patency (TLPP) and access circuit primary patency (ACPP) in hemodialysis patients with stenosed venous access.

## Methodology

### Literature Search

The electronic literature search was conducted on EMBASE, Scopus, PubMed and Web of Science for RCTs from inception to 13^th^ November 2020 using a search string [**Supplementary Table 1**], in line with the Preferred Reporting Items for Systematic Reviews and Meta-Analyses and Cochrane Guidelines (PRISMA) for IPD and network meta-analysis (NMA) [22, 23]. Only full-text articles were included. If the same publication appeared more than once, the most recent publication was used for the analysis.

We included RCTs reporting TLPP and ACPP between DCB and PTA that provided Kaplan-Meier curves. The exclusion criteria for this study were: (a) Studies only comparing differences within a single approach. (b) Studies with combination approaches including multiple intra-arterial approaches.

For studies with multiple treatment arms, we only analysed the treatment arms in our inclusion criteria. The abstracts were reviewed by 3 investigators J.J.Z., K.Y.F. and E.T. and conflicts resolved by C.W.T. The data was extracted by K.Y.F., J.J.Z. and E.T. using predefined data fields including study characteristics, patient demographics and primary endpoints.

### Risk of Bias Assessment

The RCTs were assessed for risk of bias by K.Y.F. and J.J.Z. using the Cochrane Risk of bias (RoB 2) tool for RCTs [24]. Studies were also assessed for trustworthiness and relevance with the Joanna Briggs Institute (JBI) critical appraisal tool for RCTs [25].

### Statistical Analysis

#### Reconstruction of Individual Patient Data

Prior to meta-analyses, we reconstructed IPD from published survival curves using a graphical reconstructive algorithm by Guyot et al [18]. Images of Kaplan-Meier curves from included studies were digitised to obtain the step function values and timings of the steps. Survival information of individual patients were then recovered based on the numerical solutions to the inverted Kaplan-Meier product-limit equations [18]. The IPD dataset was reconstructed by J.J.Z and K.Y.F. and were approved by E.T. by visual comparisons against original curves, and comparing reconstructed raw hazard ratio (HR) and P-values to reported values [**Supplementary Table 2, Supplementary Comparisons 1, 2**].

#### One-Stage Meta-Analysis

The Kaplan-Meier method was used to determine TLPP and ACPP for both groups. To account for between-study heterogeneity, Cox-models with random-effects gamma-frailties and stratification were conducted to determine HRs of patients undergoing DCB versus PTA. The primary analysis was based on a shared-frailty approach which assumes that individual patients are at an equal risk of death as other individuals within the same study. Then, we stratified the Cox model by study subgroups, modelling inter-study heterogeneity by assuming a baseline hazard amongst patients in each unique study.

For Cox-based models, we verified the proportional hazards (PH) assumption by examining scaled Schoenfeld residuals [26] and through the quantitative Grambsch-Therneau test [27]. Where the PH assumption was violated, we analysed restricted mean survival times (RMST) and modelled patency outcomes at prespecified epochs up to 1-, 2- and 3-years [28]. RMST is an alternative treatment outcome measure that can be estimated as the area under the survival curve up to a prespecified time horizon and hence can account for all survival information before that time horizon [28, 29]. To fully capture the dynamic changes of RMST differences and ratios, we computed RMST values over a range of values to the restriction time (tau) to trace out an evolving treatment effect profile over time [30].

Patency rates were calculated from the one-stage meta-analysis along with Greenwood 95% confidence intervals to account for censorship status. To account for the group of patients who no longer contribute to excess hazard (as seen by long plateaus on Kaplan-Meier curves), we fitted flexible parametric cure models to estimate “long term patency rates” using an identity link [31].

#### Two-Stage Meta-Analysis

We computed summary HRs for individual studies based on the reconstructed individual patient dataset and pooled them under a conventional two-stage Frequentist meta-analysis. The random-effects model was chosen in light of the high degree of heterogeneity found (*I*^*2*^). *I*^*2*^ can be interpreted as the disparity in study results due to between-study variability. In light of the potentially high degree of clinical heterogeneity of included studies (as evidenced by differences in inclusion criteria, paclitaxel dose concentrations, proportions of AVFs versus AVGs and devices utilised), a random-effects model was used. Quantitative analysis of funnel plot asymmetry was done using Egger’s regression test [32]. Heterogeneity was considered low, moderate, or considerable for *I*^*2*^ values <40%, 40-75%, and >75% respectively [33, 34]. Where considerable heterogeneity was found, we searched for extreme effect sizes (outliers) using outlier and influence analysis [35, 36]. Where outliers were consistently detected across these analyses, sensitivity analyses excluding those studies were conducted.

#### Frequentist Network Meta-Analysis

Where concentration of paclitaxel was provided, natural log-transformed hazard ratios were estimated for each IPD study and were pooled together in an NMA within a Frequentist setting. Treatment strategies were ranked using P-Scores, with higher P-Scores corresponding to greater efficacy [37].

#### Meta-Regression

To identify potential sources of heterogeneity, we performed a meta-regression with aggregate level data. Prior to meta-regression, we imputed missing means and standard deviations (SDs) from medians, ranges (minimum to maximum), and interquartile ranges (IQRs) using the methods proposed by Hozo et al [38], Wan et al [39] and Furukawa et al [40]. Conventional mixed effects meta-regression of variables (with more than 10 studies) were performed against logarithmic transformed hazard ratios.

All analyses were conducted in R-4.0.0 (with packages ‘digitize’, ‘survival’, ‘flexsurvcure’, ‘metafor’. ‘dmetar’ and ‘netmeta’). P < 0.05 were regarded to indicate statistical significance.

## Results

### Study Selection

The search strategy retrieved 503 studies; after de-duplication and screening, 11 RCTs [16, 17, 41-48] comprising 1,243 patients were eligible and included in the meta-analysis [**Figure 1**]. One RCT was excluded as no curve was provided for graphical reconstruction to retrieve patient-level patency information [49].

**Figure 1.**
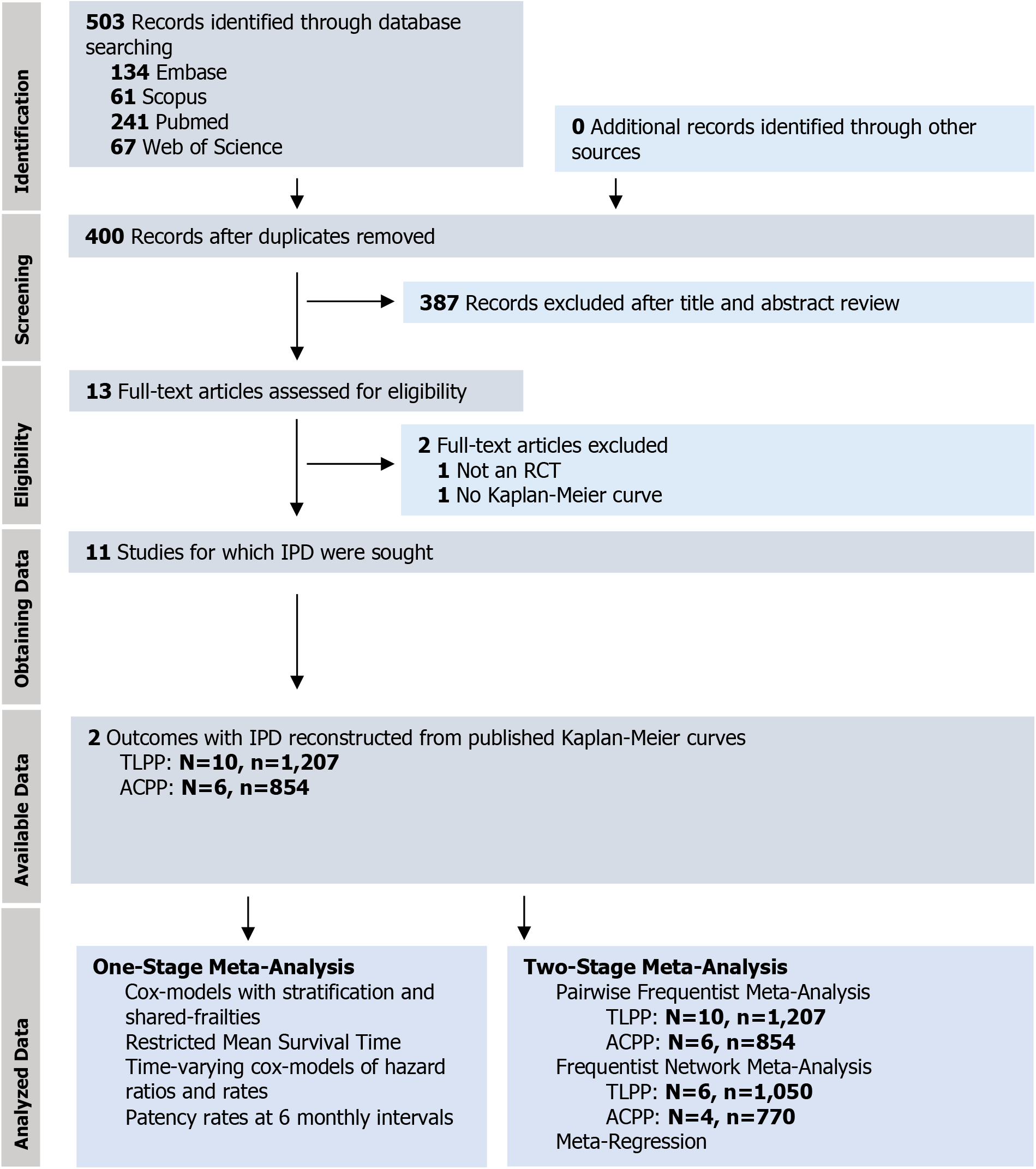
PRISMA flowchart. PRISMA indicates preferred reporting items for systematic reviews and meta-analyses. N=, number of studies; n=, number of patients; TLPP, target lesion primary patency; ACPP, access circuit primary patency; IPD, individual patient data; RCT, randomized controlled trials.

### Study Characteristics

Breakdown of studies and extracted outcomes are reported in **Table 1** and in **Supplementary Table 2** respectively. Of note: 1) 5 of the 11 studies had 20 or less patients per arm; 2) all studies had a higher percentage of males except Liao et al [47]; 3) a significant proportion of patients in most studies received antiplatelet treatment, either preoperatively or postoperatively; 4) 6 studies reported a significant benefit with DCB based on primary and secondary endpoints, while the other 5 found no significant difference compared with PTA; 5) 4 studies involved AVGs, of which 1 study (Liao et al [47]) studied only AVGs; the other 7 involved only AVFs.

**Table 1A.**
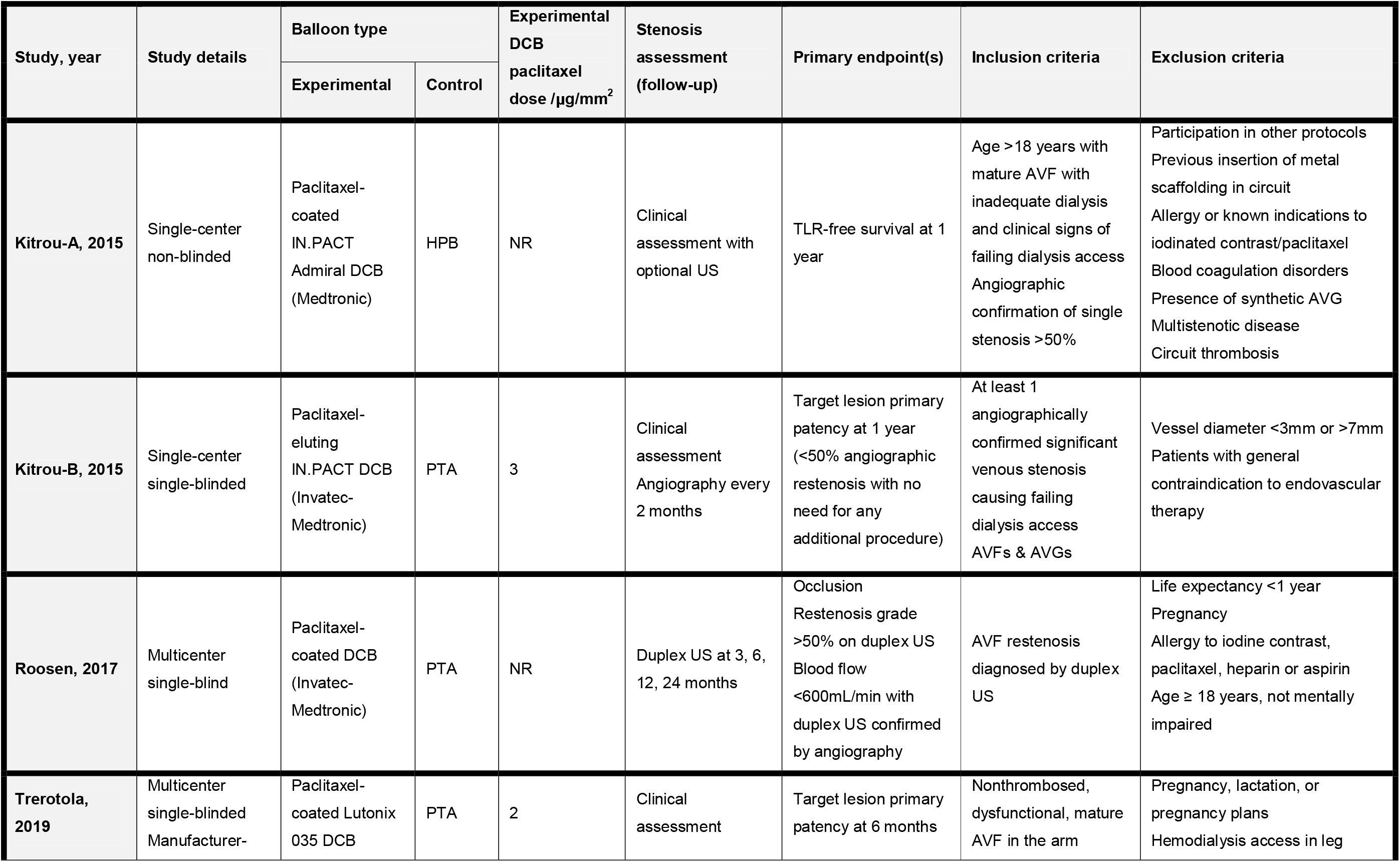
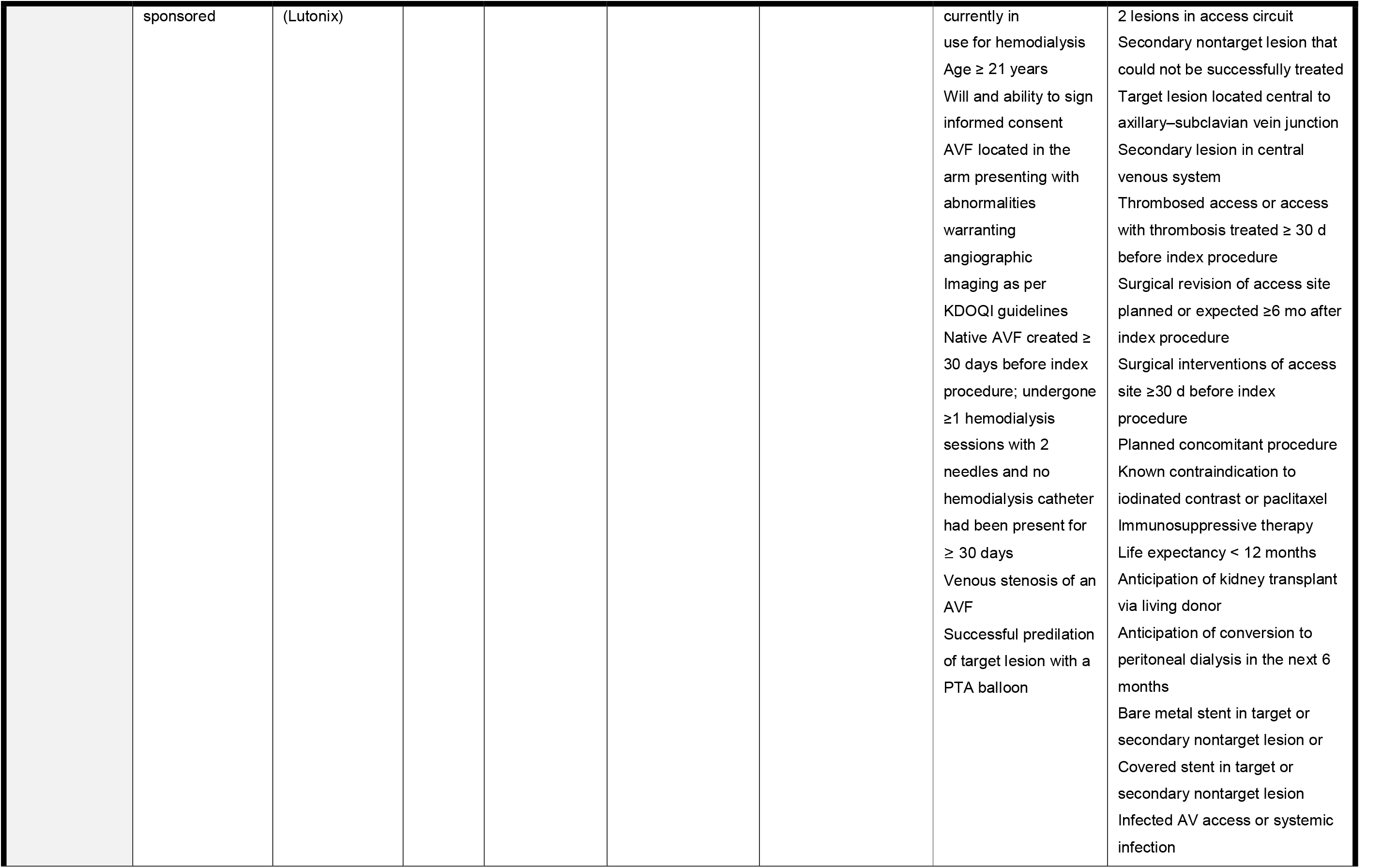
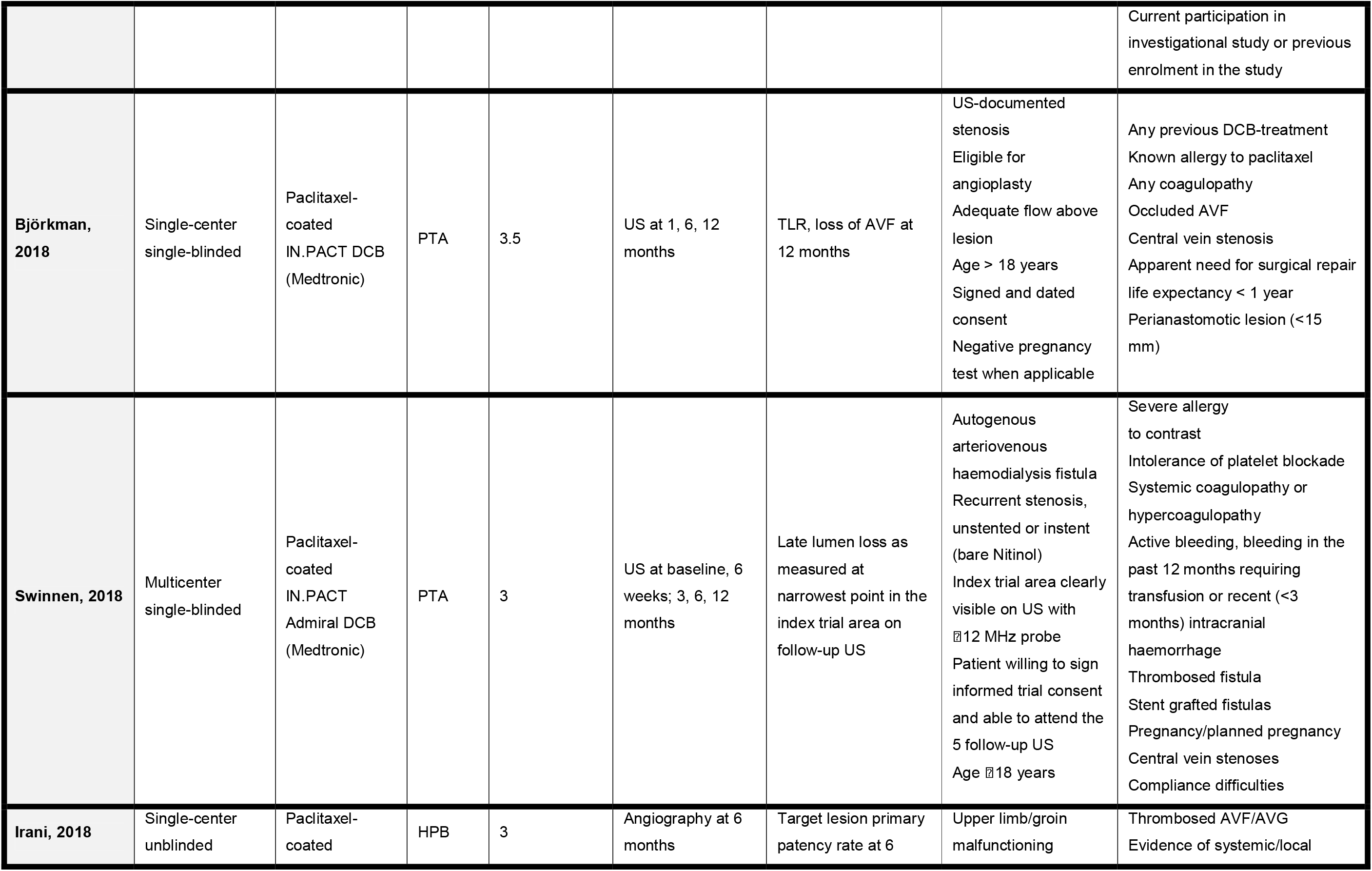
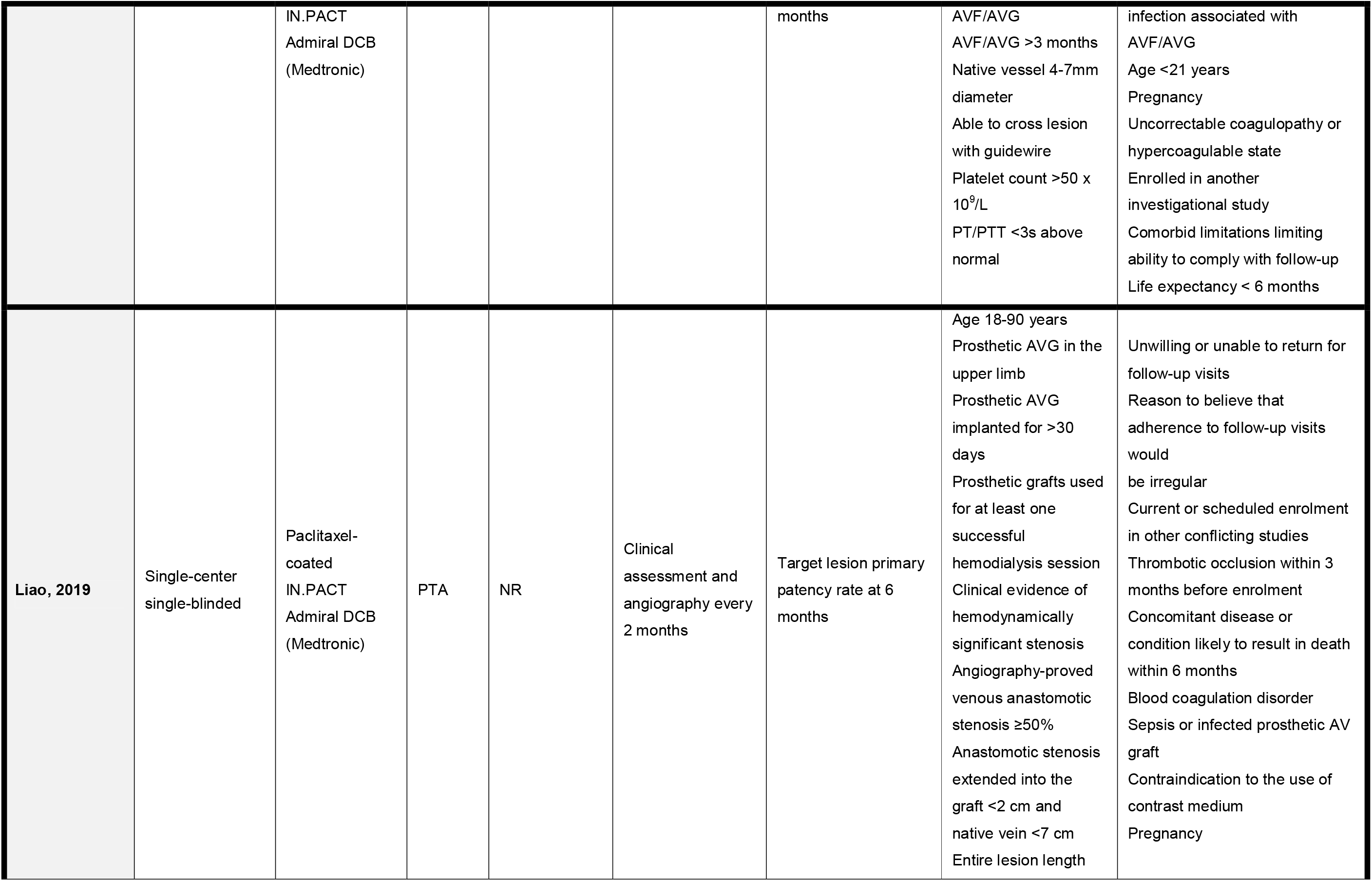
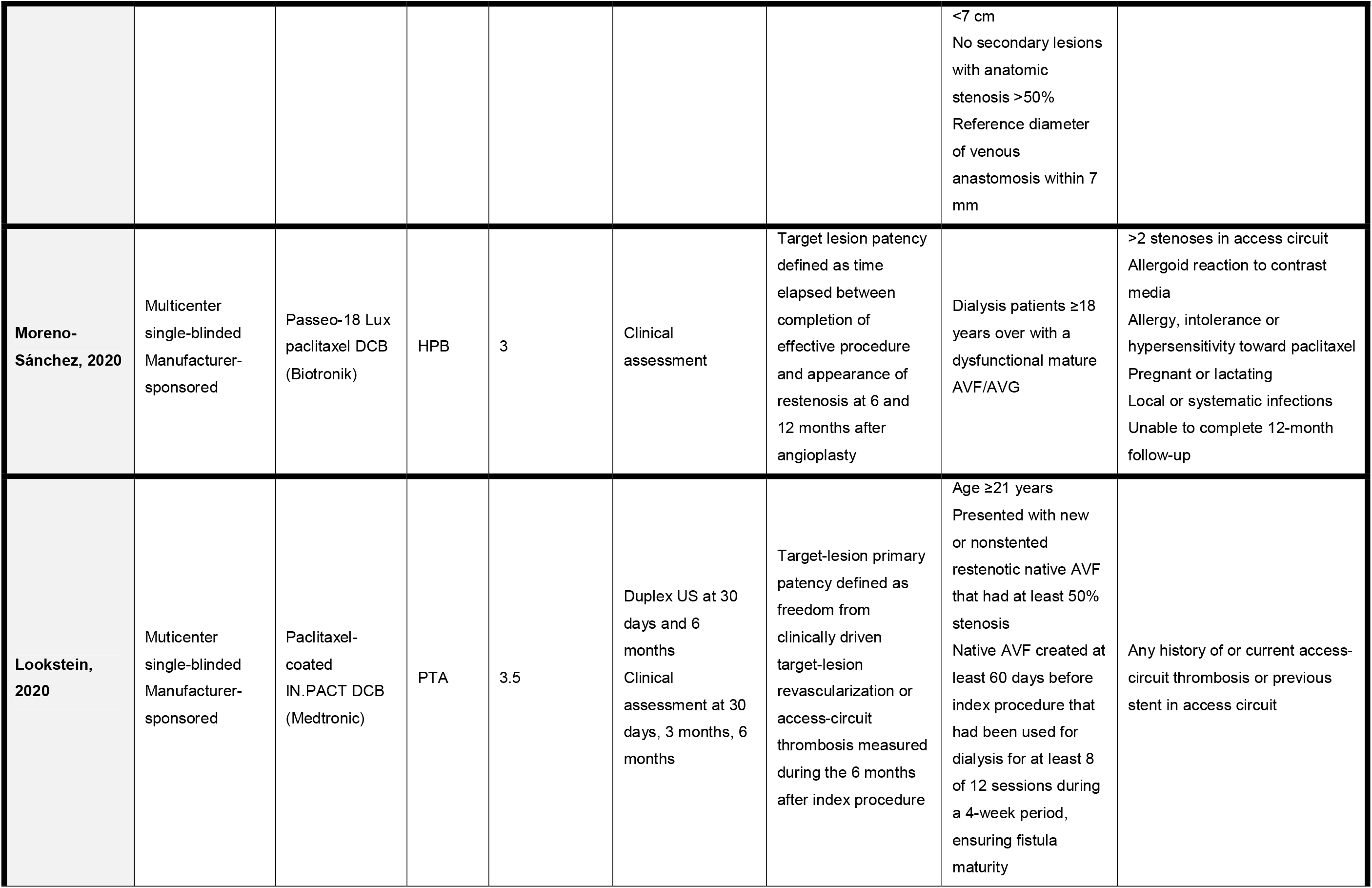
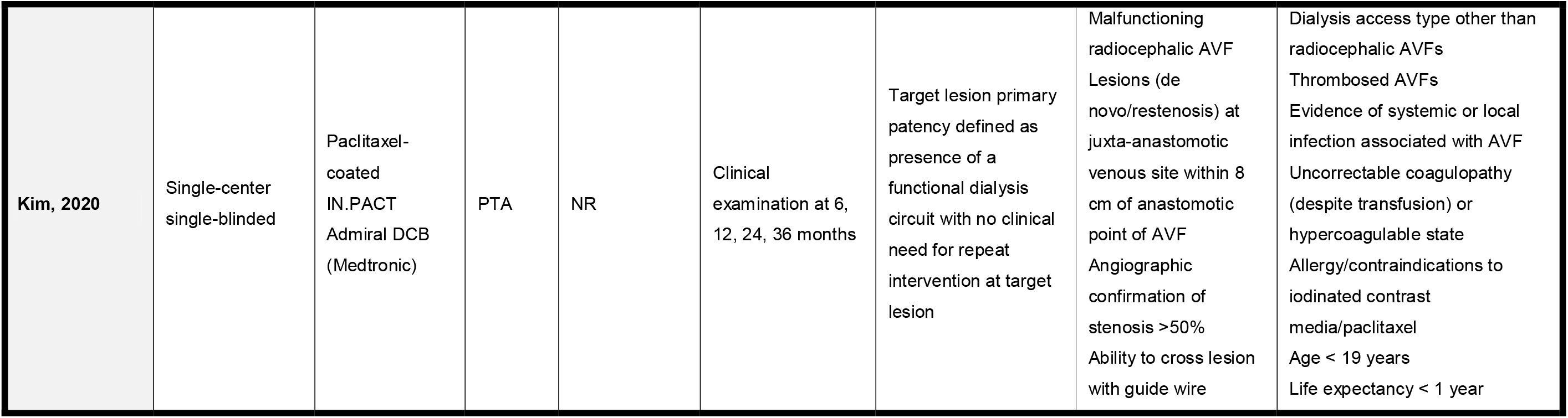
Characteristics of study designs.

**Table 1B.** Patient characteristics of included studies.

| Study, year | No. of patients (male, female) |  | Age |  | Baseline dialysis access age |  | Follow-up period (months) | Diabetes mellitus (%) |  | Hypertension (%) |  | AVFs (%) |  |
| --- | --- | --- | --- | --- | --- | --- | --- | --- | --- | --- | --- | --- | --- |
|  | DCB | PTA | DCB | PTA | DCB | PTA |  | DCB | PTA | DCB | PTA | DCB | PTA |
| **Kitrou-A, 2015** | 20 (12M, 8F) | 20 (14M, 6F) | 64.3±14.5 | 57±14.2 | 2.13±2.12 years | 2.74±2.4 years | 12 | 20 | 35 | 15 | 15 | 100 | 100 |
| **Kitrou-B, 2015** | 20 (15M, 5F) | 20 (14M, 6F) | 65.7±13.2 | 62.5±15.4 | 2.5±2.0 years | 2.5±3.2 years | 12 | 20 | 20 | 15 | 10 | 35 | 35 |
| **Roosen, 2017** | 16 (7M, 9F) | 18 (14M, 4F) | 80 [71-86] | 83 [78-86] | NR | NR | 24 | 37.5 | 22.2 | 43.8 | 38.9 | 83.5 | 83.3 |
| **Trerotola, 2019** | 141 (87M, 54F) | 144 (85M, 59F) | 64±15 | 61±13 | 1164±973 days | 1056±1122 days | 24 | 58.2 | 65.3 | 94.3 | 98.6 | 100 | 100 |
| **Björkman, 2018** | 18 (10M, 8F) | 18 (13M, 5F) | 67.4 (46-87) | 67.0 (28-82) | 165 (45-1165) days | 292 (80-3800) days | 12 | 61.1 | 61.1 | 77.8 | 88.9 | 100 | 100 |
| **Swinnen, 2018** | 68 (42M, 26F) | 60 (37M, 23F) | 65.2±13.6 | 64.5±13.9 | 2 ≤ 6 months<br>66 > 6 months | 3 ≤ 6 months<br>57 > 6 months | 12 | 41.2 | 18.3 | 17.6 | 10 | 100 | 100 |
| **Irani, 2018** | 59 (39M, 20F) | 60 (40M, 20F) | 59.0±11.5 | 59.4±8.80 | 44.4±58.6 months | 47.3±54.3 months | 12 | 62.7 | 56.7 | 93.2 | 91.7 | 88.1 | 76.7 |
| **Liao, 2019** | 22 (3M, 19F) | 22 (9M, 11F) | 70.4±10.6 | 65.9±15.9 | 28 [15-41] months | 34 [18-68] months | 12 | 77.3 | 54.5 | 81.8 | 50 | 0 | 0 |
| **Moreno-Sánchez, 2020** | 70 (55M, 15F) | 78 (52M, 26F) | 69±12.99 | 71±11.31 | All mature AVF/AVG; exact figures not specified |  | 12 | 28.5 | 23 | 65.7 | 70.5 | 100 | 100 |
| **Lookstein, 2020** | 170 (112M, 58F) | 160 (101M, 59F) | 65.8±13.1 | 65.5±13.4 | All ≥60 days; exact figures not specified |  | 6 | 62.9 | 68.8 | 91.2 | 94.4 | 100 | 100 |
| **Kim, 2020** | 20 (12M, 8F) | 19 (9M, 10F) | 60.7±12.2 | 63.7±11.8 | 27.3±30.7 months | 19.1±11.1 months | 36 | 80 | 78.9 | NR | NR | 100 | 100 |

Values in square brackets are interquartile range, values in circular brackets are range. DCB, drug coated balloon; PTA, percutaneous transluminal angioplasty; HPB, high pressure balloon; TLR, target lesion restenosis; TLPP, target lesion primary patency; ACPP, access circuit primary patency; AVF, arteriovenous fistula; AVG, arteriovenous graft; US, ultrasound; KDOQI, Kidney Disease Outcomes Quality Initiative; PT, prothrombin time; PTT, partial thromboplastin time; NR, not reported

### Reconstructed Curves

All included studies had IPD reconstructed from published Kaplan-Meier curves [**Figure 2**]. Comparisons against original Kaplan-Meier curves are shown in **Supplementary Comparisons 1 & 2**. The reconstructed dataset is available on request.

**Figure 2.**
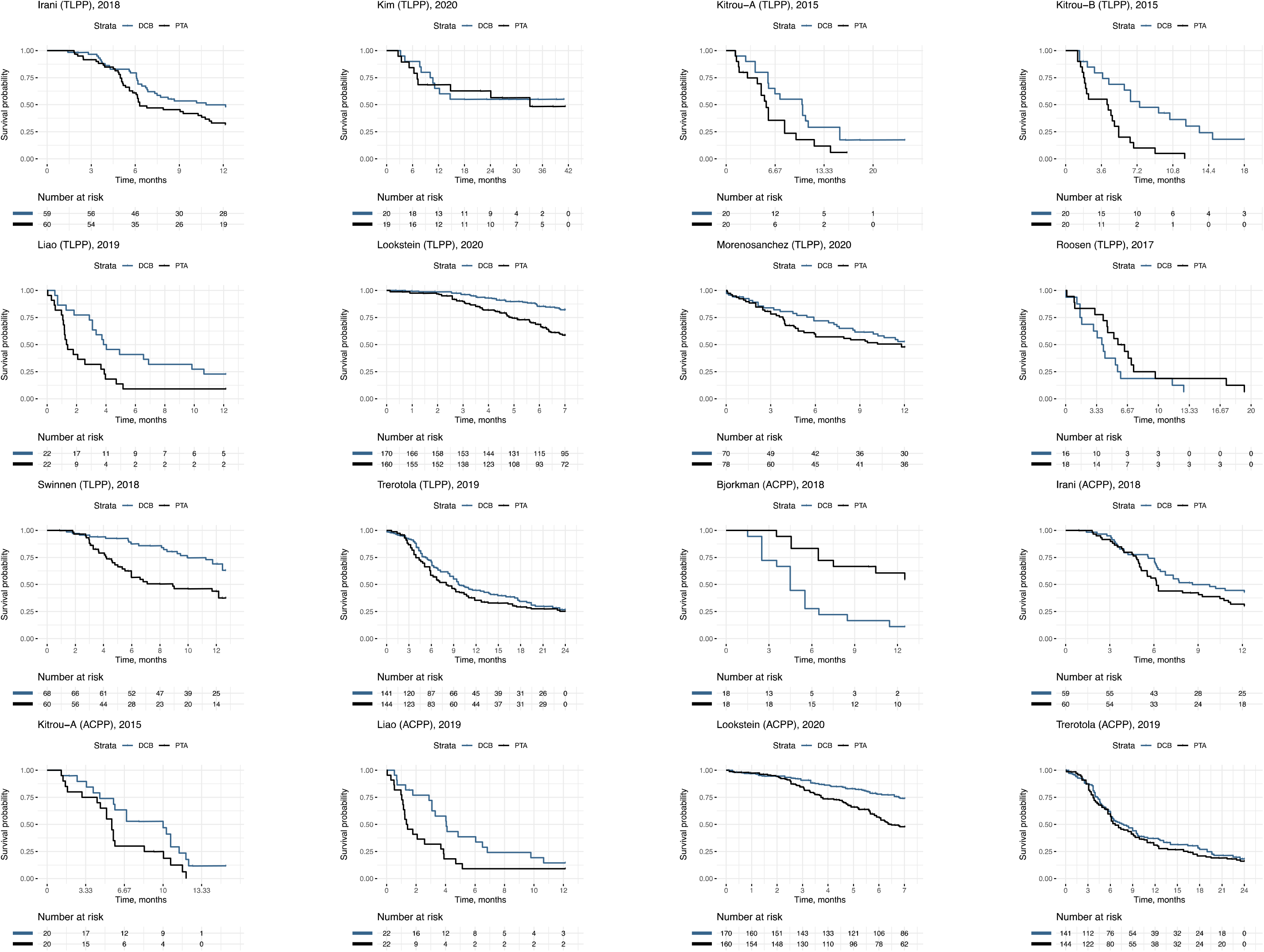
Reconstructed Curves. DCB, drug coated balloon; PTA, percutaneous transluminal angioplasty.

### Quality Assessment of Trials

Our risk-of-bias analysis [**Table 2**] yielded overall ‘some concerns’ for all 11 studies. As all studies were single-blinded or non-blinded, they ran the risk of observer bias, wherein knowledge of which patients were assigned DCB or PTA might have influenced the judging of outcomes. Particularly, decisions on the need for reintervention in all studies may have been influenced by knowledge of assignment to treatment or control arm, since the reinterventions in most studies involved clinical evaluation of graft dysfunction. Our JBI analysis [**Supplementary Table 3**] noted that Björkman et al [44] discontinued their trial after only 39 patients were recruited, despite requiring a sample size of 140 in their initial power calculations.

**Table 2.** Quality Assessment of Trials: Cochrane risk of bias (ROB) assessment of randomized controlled trials.

| Cochrane Risk-of-bias tool for randomized controlled trials (Consensus) |  |  |  |  |  |  |  |
| --- | --- | --- | --- | --- | --- | --- | --- |
| Study ID | Outcome | Randomization process | Deviations from intended interventions | Missing outcome data | Measurement of the outcome | Selection of the reported result | Overall |
| Kitrou-A, 2015 | TLR-free survival | + | + | + | - | + | ! |
| Kitrou-B, 2015 | TLPP | + | + | + | - | + | ! |
| Roosen, 2017 | Occlusion Restenosis Blood flow measurement | ! | + | + | - | + | ! |
| Trerotola, 2019 | TLPP | + | + | + | - | + | ! |
| Björkman, 2018 | TLR | + | + | + | - | ! | ! |
| Swinnen, 2018 | Late lumen loss | + | + | + | - | + | ! |
| Irani, 2018 | TLPP | + | + | + | - | + | ! |
| Liao, 2019 | TLPP | + | + | + | - | + | ! |
| Moreno-Sánchez, 2020 | TLPP | + | + | + | - | + | ! |
| Lookstein, 2020 | TLPP | + | + | + | - | + | ! |
| Kim, 2020 | TLPP | + | + | + | - | + | ! |

TLR, target lesion restenosis; TLPP, target lesion primary patency

### Survival Outcomes

#### Target Lesion Primary Patency

Among 10 RCTs comprising 1,207 patients, the stratified and gamma-frailty models yielded HRs in favour of DCB (shared-frailty HR=0.62, 95%-CI: 0.53–0.73, P<0.001; stratified HR=0.62, 95%-CI: 0.53–0.73, P<0.001) [**Figure 3**]. Patency rates at intervals of 6-months are reported in **Figure 3**.

**Figure 3.**
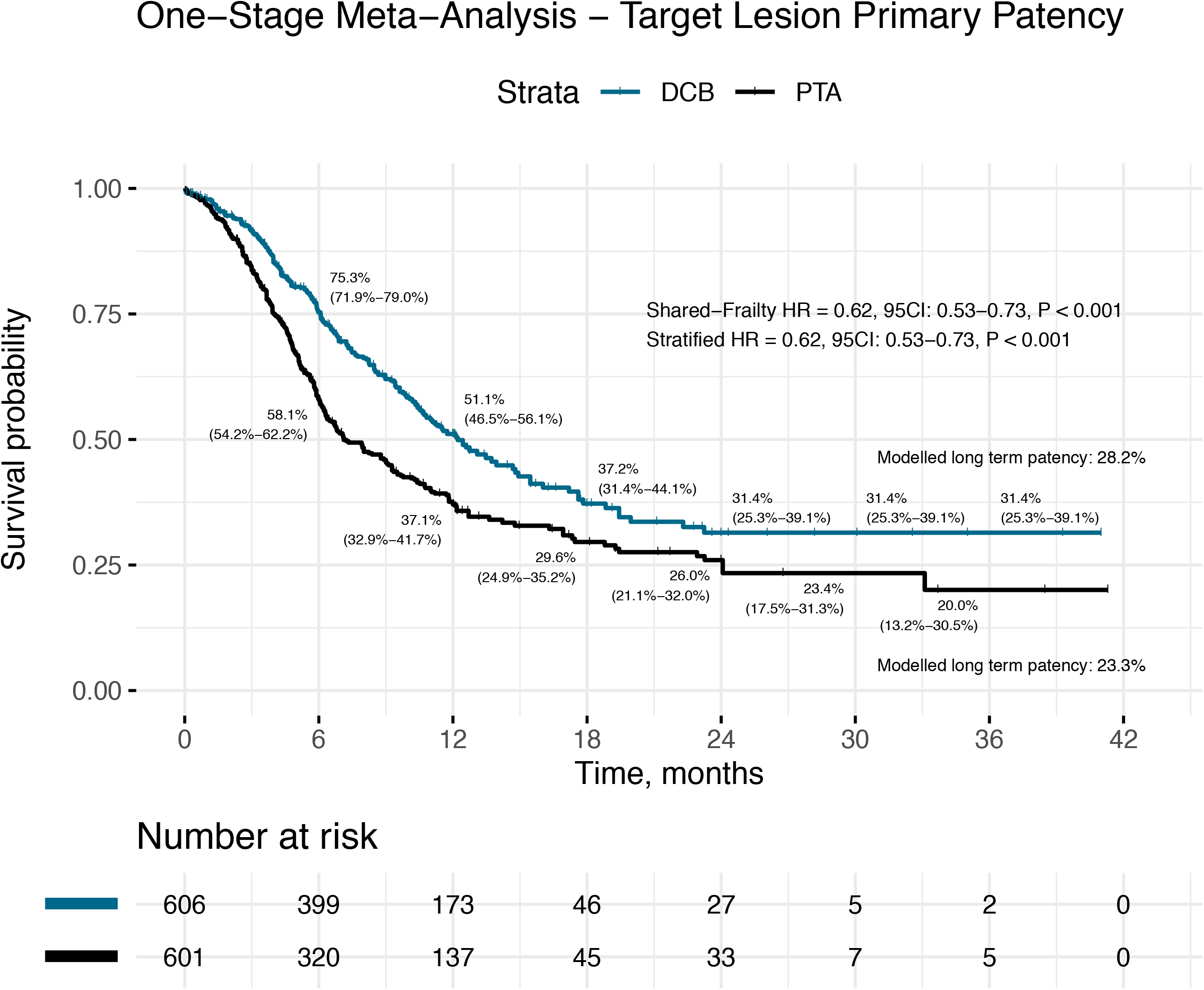
Kaplan-Meier plot and number-at-risk table for target lesion primary patency. HR, hazard ratio; 95CI, 95% confidence intervals; DCB, drug coated balloon; PTA, percutaneous transluminal angioplasty.

While time-varying effects were noted (Grambsch-Therneau test, P=0.005) [**Supplementary Figure 1A**], dynamic RMST curves demonstrate that DCB is consistently favoured throughout the follow-up period [**Figure 4A, B**].

**Figure 4.**
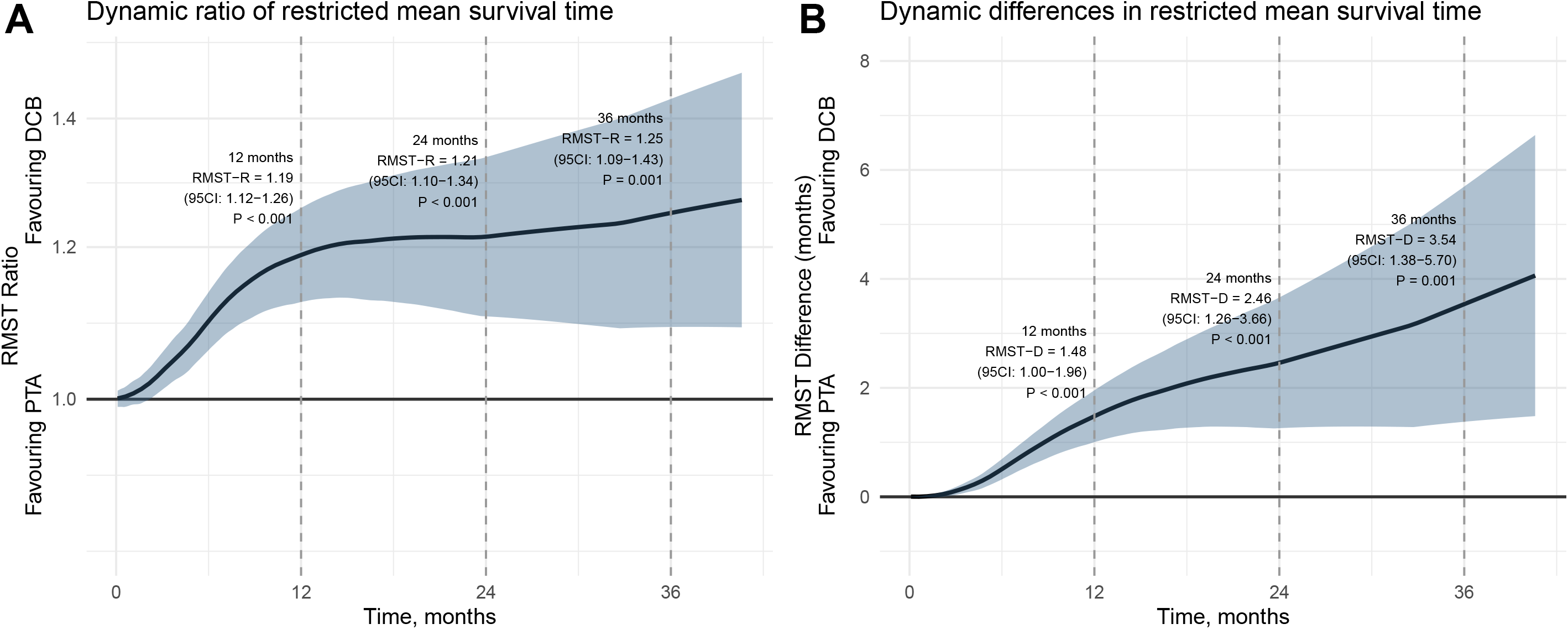
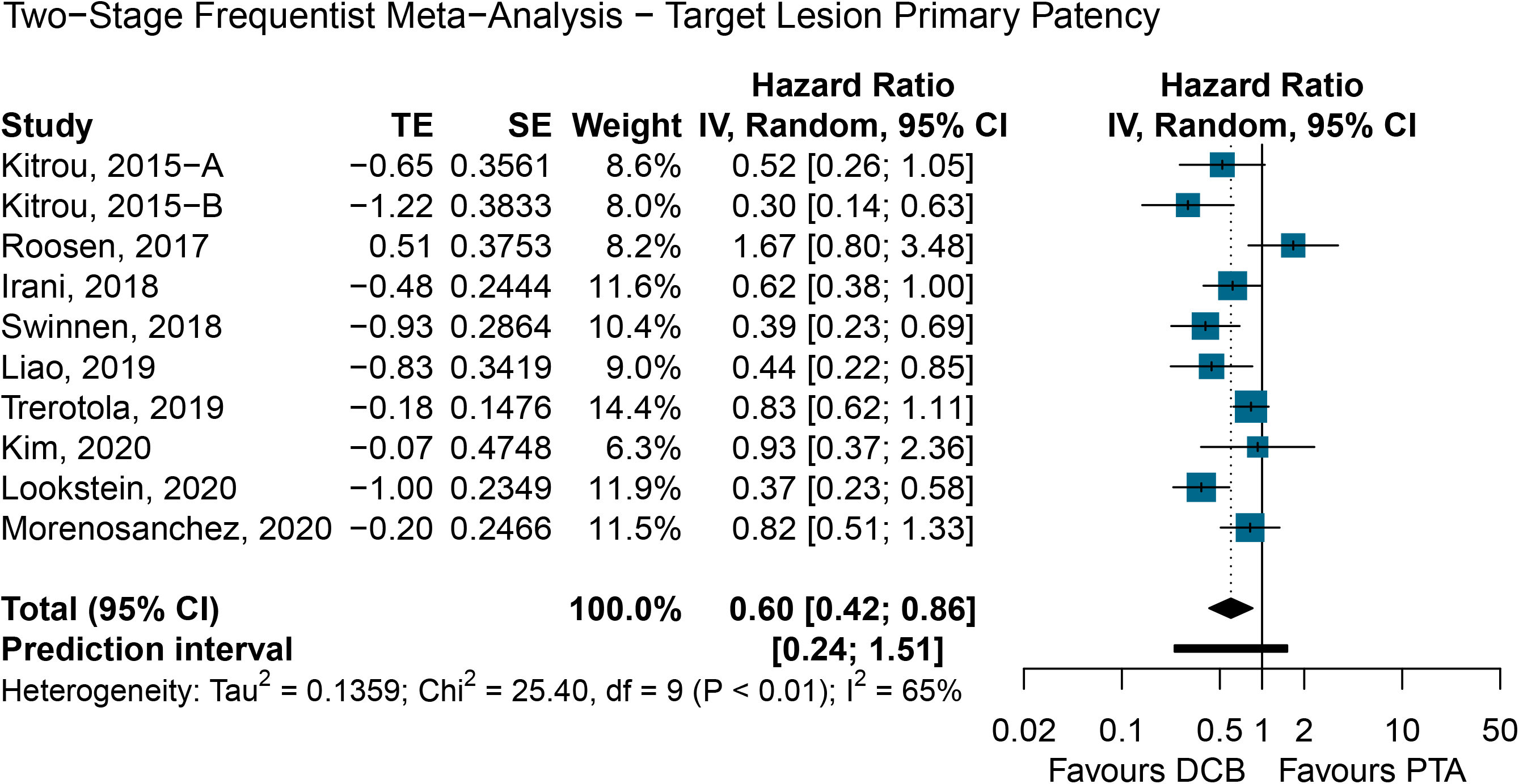
(A) Dynamic RMST Ratios (B) Dynamic RMST Differences for Target Lesion Primary Patency (C) Two-stage Frequentist meta-analysis for target lesion primary patency. RMST-R, restricted mean survival time ratio; RMST-D, restricted mean survival time difference; HR, hazard ratio; 95CI, 95% confidence intervals; DCB, drug coated balloon; PTA, percutaneous transluminal angioplasty.

This was consistent within a two-stage Frequentist random-effects model (HR=0.60, 95%-CI: 0.42–0.86, P=0.018, *I*^*2*^=65%) [**Figure 4C**]. The funnel plot was visually symmetrical and did not suggest publication bias (Egger’s test = 0.507) [**Supplementary Figure 4A**].

#### Access Circuit Primary Patency

Among 6 RCTs comprising 854 patients, the stratified and gamma-frailty models yielded HRs in favour of DCB (shared-frailty HR=0.72, 95%-CI: 0.60–0.87, P<0.001; stratified HR=0.72, 95%-CI: 0.60–0.86, P<0.001) [**Figure 5A**]. Time-varying effects were not found (Grambsch-Therneau test, P=0.300) [**Supplementary Figure 1B**]. Patency rates at intervals of 6-months are reported in **Figure 5A**.

**Figure 5.**
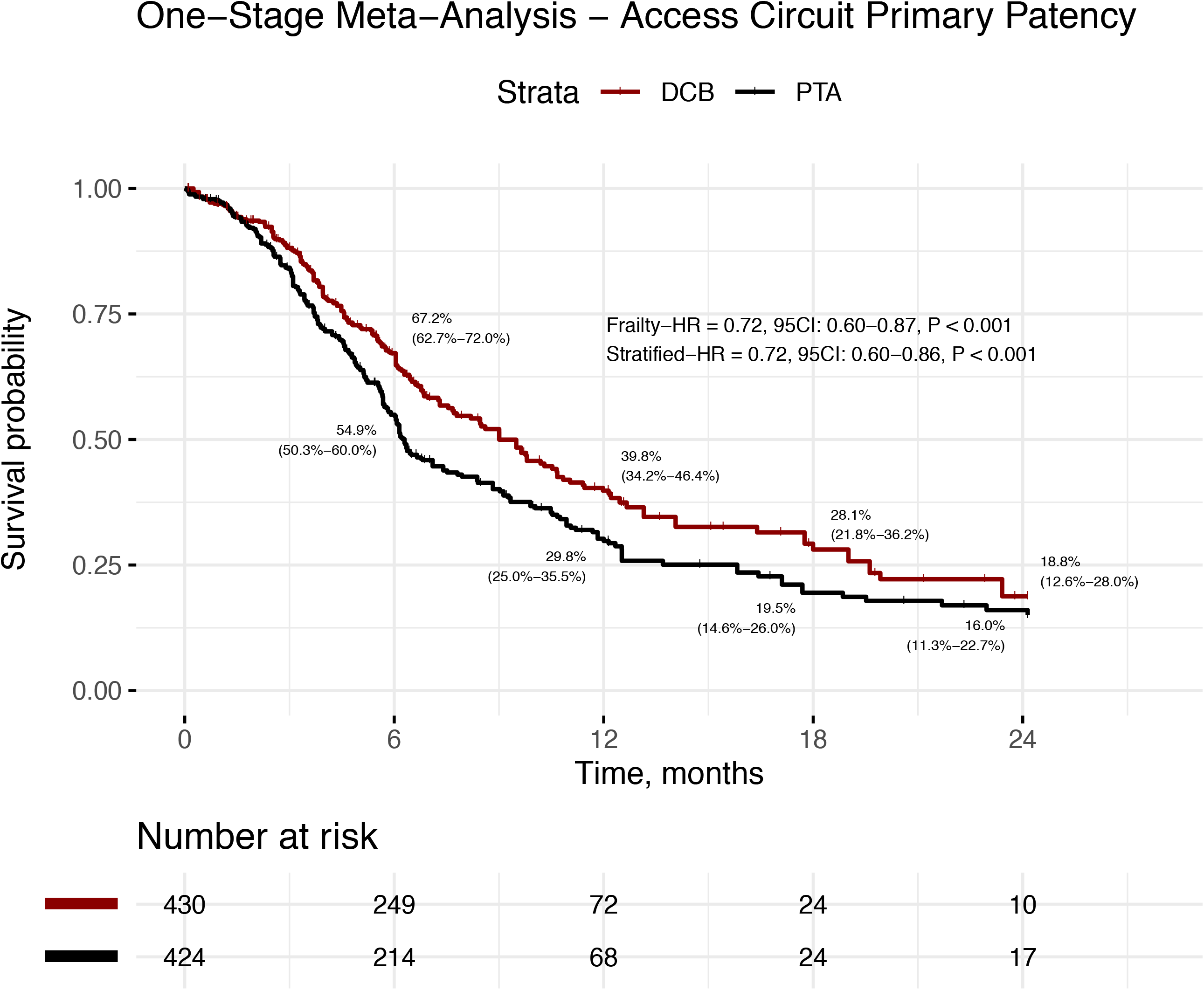
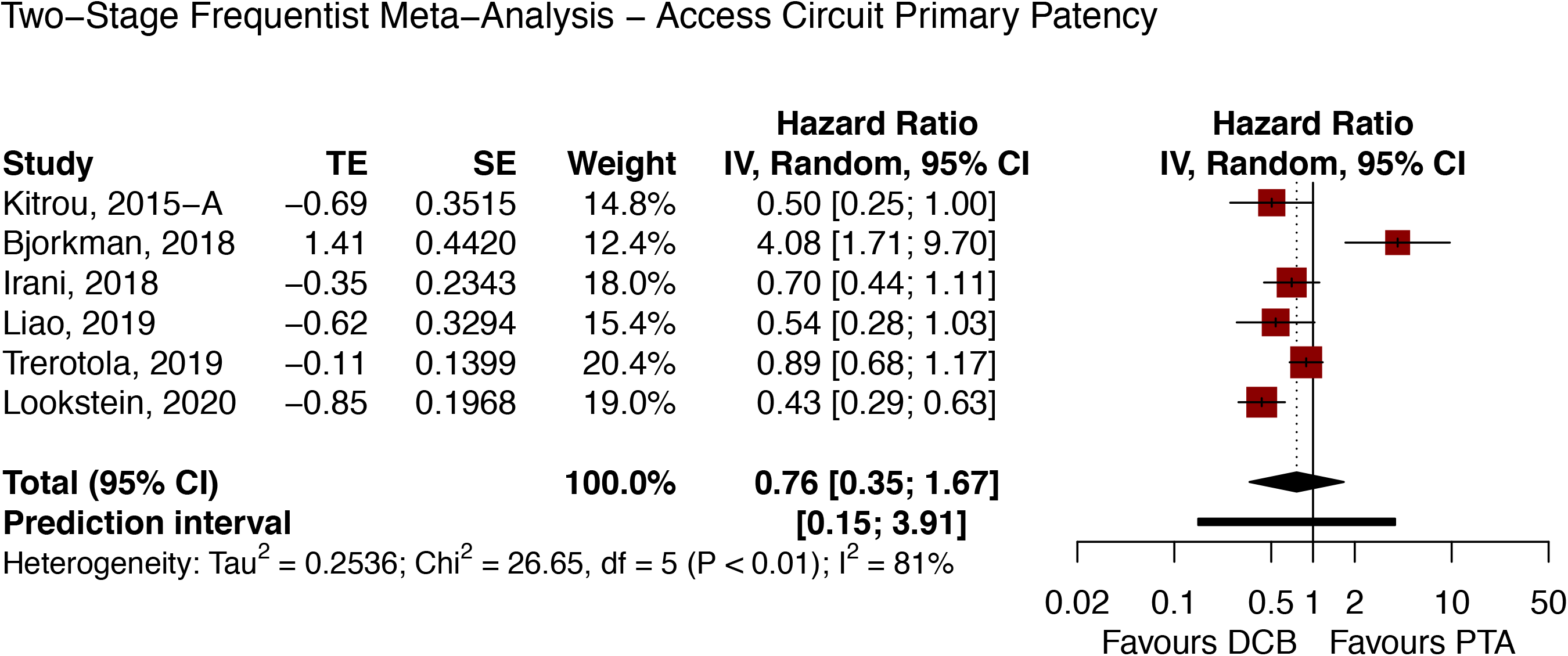
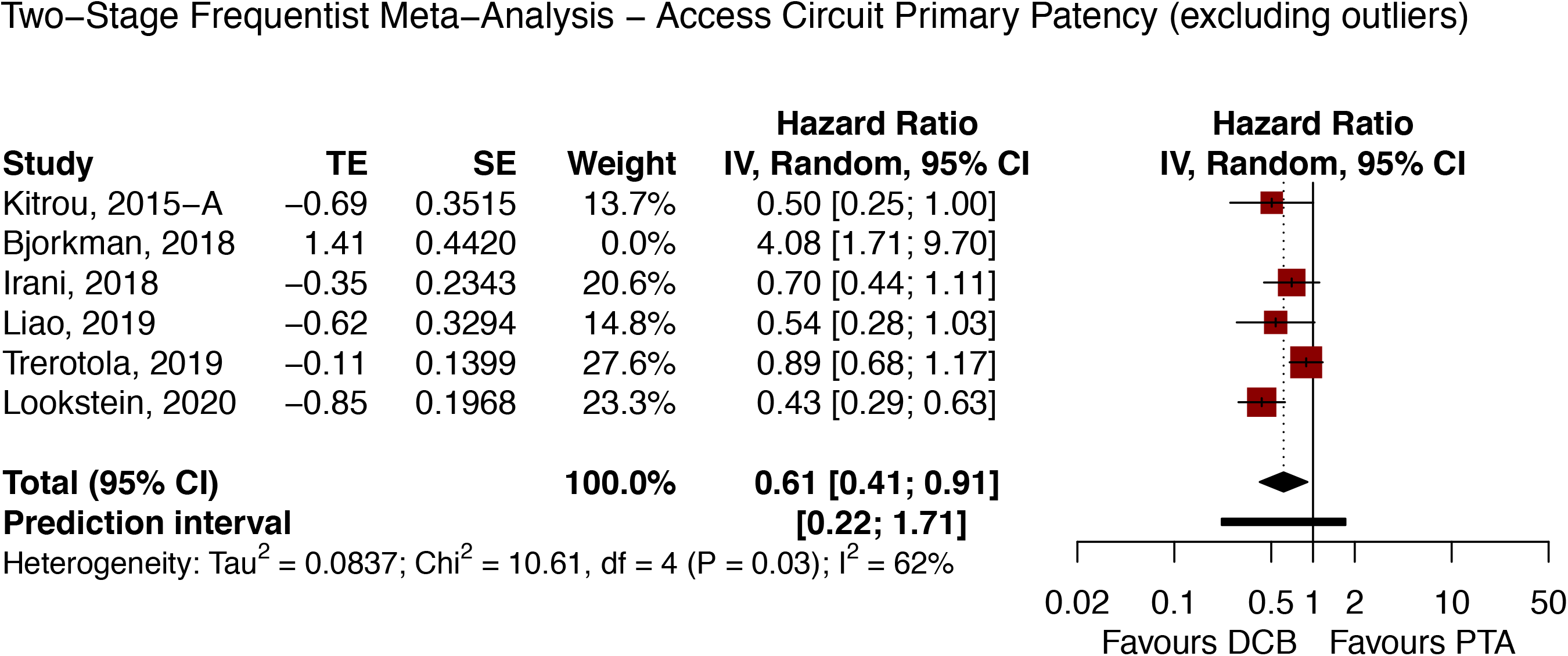
(A) Kaplan-Meier plot and number-at-risk table for access circuit primary patency (B) Two-stage Frequentist meta-analysis for access circuit primary patency (C) Two-stage Frequentist meta-analysis for access circuit primary patency excluding outliers. HR, hazard ratio; 95CI, 95% confidence intervals; DCB, drug coated balloon; PTA, percutaneous transluminal angioplasty.

This was inconsistent within the overall two-stage Frequentist random-effects model (HR=0.76, 95%-CI: 0.35–1.67, P=0.414, *I*^*2*^=81%) [**Figure 5B**]. In view of the considerable heterogeneity found, we undertook efforts to identify potential outliers. The Baujat plot suggested that Björkman et al [44] contributed to the most between-study heterogeneity [**Supplementary Figure 5A**]. At cut-offs proposed by Viechtbauer and Cheung [35], Björkman et al was likewise detected as an influential case [**Supplementary Figure 5B**]. Leave-one-out analyses showed that omission of Björkman et al yielded the least between-study heterogeneity (*I*^2^=62%) [**Supplementary Figure 5C**].

Sensitivity analysis excluding Björkman et al significantly favoured DCB (two-stage Frequentist random-effects model HR=0.61, 95%-CI: 0.41–0.91, P=0.027, *I*^*2*^=62%) [**Figure 5C**]. The one-stage model excluding this study yielded congruent results with one-stage meta-analysis of the overall cohort [**Supplementary Figure 6**]. The funnel plot was visually asymmetrical and suggestive of publication bias [**Supplementary Figure 4B**].

#### Frequentist Network Meta-Analysis of Paclitaxel Concentrations

6 RCTs comprising 1,050 patients were included in a network meta-analysis to evaluate TLPP within a random-effects Frequentist setting (*I*^*2*^=56.1%). Among all comparisons against PTA, only DCB 3.5μg/mm^2^ (HR=0.37, 95%-CI: 0.17–0.80, P=0.012) and DCB 3.0μg/mm^2^ (HR=0.52, 95%-CI: 0.34–0.80, P=0.002) had a statistically significant TLPP advantage [**Table 3**]. Indirect comparisons between DCB 3.5μg/mm^2^ and DCB 3.0μg/mm^2^ showed no differences (HR=0.71, 95%-CI: 0.29–1.70, P=0.437) [**Table 3**].

**Table 3.** Frequentist Network Meta-Analysis.

| Access Circuit Primary Patency |  |  |  |  |
| --- | --- | --- | --- | --- |
| Target Lesion Primary Patency | **DCB**<br>**(Paclitaxel 3.5µg/mm<sup>2</sup>)**<br>TLPP P-Score = **<u>0.905</u>**<br>ACPP P-Score = **<u>0.982</u>** | 1.645<br>(95%-CI: 0.903 - 2.997)<br>P = 0.104 | **2.093**<br>**(95%-CI: 1.304 - 3.359)**<br>**P = 0.002** | **2.343**<br>**(95%-CI: 1.593 - 3.446)**<br>**P < 0.001** |
|  | 0.705<br>(95%-CI: 0.292 - 1.703)<br>P = 0.437 | **DCB**<br>**(Paclitaxel 3.0µg/mm<sup>2</sup>)**<br>TLPP P-Score = 0.697<br>ACPP P-Score = 0.599 | 1.272<br>(95%-CI: 0.745 - 2.172)<br>P = 0.378 | 1.424<br>(95%-CI: 0.900 - 2.255)<br>P = 0.131 |
|  | 0.442<br>(95%-CI: 0.157 - 1.245)<br>P = 0.122 | 0.627<br>(95%-CI: 0.280 - 1.402)<br>P = 0.255 | **DCB**<br>**(Paclitaxel 2.0µg/mm<sup>2</sup>)**<br>TLPP P-Score = 0.295<br>ACPP P-Score = 0.327 | 1.120<br>(95%-CI: 0.851 - 1.473)<br>P = 0.419 |
|  | **0.369**<br>**(95%-CI: 0.170 - 0.800)**<br>**P = 0.012** | **0.523**<br>**(95%-CI: 0.344 - 0.795)**<br>**P = 0.002** | 0.834<br>(95%-CI: 0.419 - 1.659)<br>P = 0.605 | **PTA**<br>TLPP P-Score = 0.103<br>ACPP P-Score = 0.092 |

Treatment effects are described in hazard ratios. HR, hazard ratio; 95%-CI, 95% confidence intervals; DCB, drug coated balloon; PTA, percutaneous transluminal angioplasty, TLPP, target lesion primary patency; ACPP, access circuit primary patency

3 RCTs comprising 736 patients were included in a network meta-analysis to evaluate ACPP within a random-effects Frequentist setting. Heterogeneity was not estimable. Björkman et al was excluded from this analysis in view of the upstream influence analysis. Among all comparisons against PTA, only DCB 3.5μg/mm^2^ (HR=2.34, 95%-CI: 1.59–3.45, P<0.001) had a statistically significant ACPP advantage [**Table 3**]. Indirect comparisons between DCB 3.5μg/mm^2^ and DCB 2.0μg/mm^2^ was significantly in favour of DCB 3.5μg/mm^2^ (HR=2.09, 95%-CI: 1.30–3.36, P=0.002) [**Table 3**].

The comparison adjusted funnel plots were symmetrical [**Supplementary Figure 7**]. Within the network meta-analysis, we surmise that the transitivity assumption was likely to be met, based on the observations that the common treatment (PTA) was reasonably consistent across trials, effect modifiers were equally distributed across studies, and participants may, in principle, be randomized to any of the treatments being compared in the network. Given the star shaped nature of our network of treatment arms, statistical inconsistency could not be evaluated [**Supplementary Figure 8**].

#### Meta-Regression

Meta-regression of proportions of males, diabetics, arteriovenous fistulas and publication year against logarithmic transformed HRs for TLPP did not demonstrate any significant associations [**Table 4**]. Meta-regression for variables against ACPP between DCB and PTA was not conducted as there were insufficient studies.

**Table 4.**
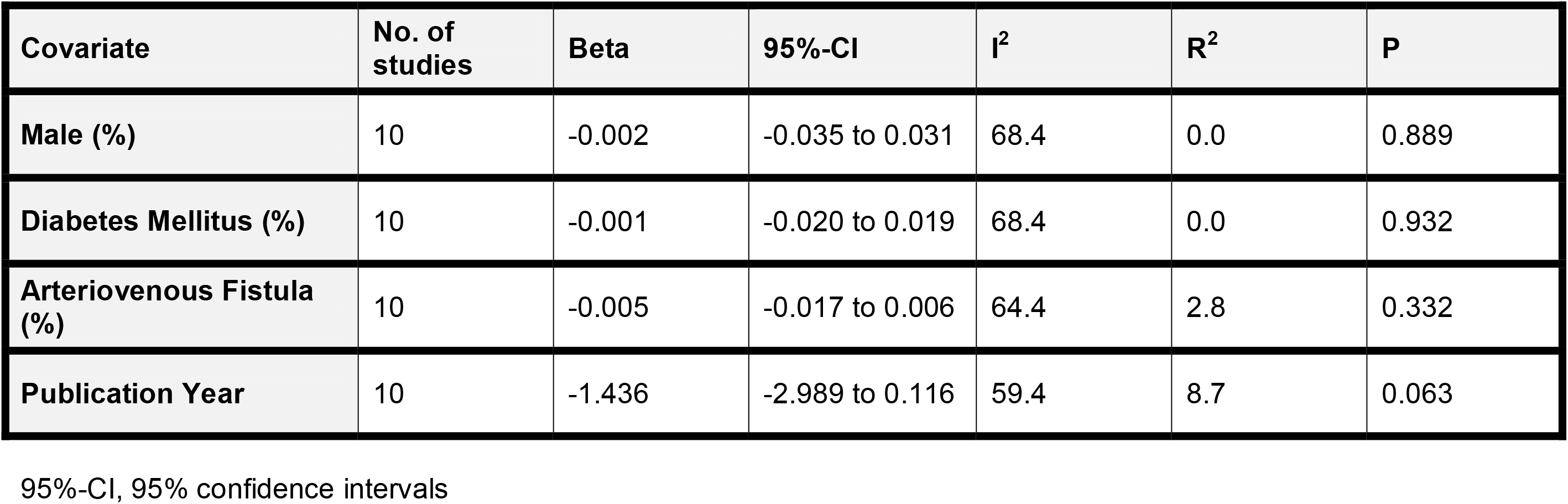
Two-Stage Meta-Regression (Target Lesion Primary Patency)

## Discussion

In view of cost [41, 50], potential morbidity [51] and uncertain theoretical benefit in prolonging patency rates, DCB angioplasty has yet to earn its place as the “holy grail” for dysfunctional hemodialysis venous access [52]. In 2019, in a meta-analysis of RCTs and retrospective studies by Wee et al [12], DCB was concluded to be superior to PTA (6-month TLPP RR=0.57, 95%-CI: 0.44-0.74, P<0.0001). In contrast, in a 2020 meta-analysis by Liao et al [14], DCB did not demonstrate significant patency benefit (6-month TLPP RR=0.75, 95%-CI: 0.56-1.01, P=0.06), even in the AVF-only subgroup.

Between the randomized trials conducted, significant heterogeneity was quantitatively, and qualitatively noted with regard to patient recruitment and access details – rendering the interpretation of its authentic efficacy challenging. Devices utilized (from IN.PACT Admiral, Passeo-Lux and Lutonix) varied in a multitude of aspects; such as its excipient, a chemical added to locally retain paclitaxel at the intended site of action.

Amidst this unresolved backdrop, this patient-level meta-analysis of 11 RCTs comprising 1,243 patients presents a statistically robust and up-to-date pool of evidence demonstrating a consistent benefit of using DCB over PTA in prolonging TLPP.

Despite incorporating random-effects shared-frailties and stratification to account for clinical heterogeneity within the one-stage model, patients treated with DCB were at a lower hazard rate of restenosis. Sensitivity analyses with aggregate data within a random-effects Frequentist models likewise re-affirm the upstream findings. In view of the violation of the proportionality assumption, dynamic restricted mean survival times consistently favoured DCB across the follow-up period. Collectively, these strongly suggest that the benefit of paclitaxel is likely a reflection of a true biological phenomenon, attesting to its superiority to plain old balloon angioplasty.

Albeit the promising outcomes in prolonging TLPP, comparisons of ACPP in the overall two-stage Frequentist random-effects model showed no significant differences as opposed to the significant differences demonstrated within one-stage models. Nonetheless, sensitivity analysis excluding outliers significantly favoured DCB in both one & two-stage models.

Other factors which are not directly related to the target lesion may cause eventual dysfunction elsewhere in the circuit. Perhaps, the puncture of AVGs and AVFs during the procedure could cause platelet thrombi and cytokine release; even though in most instances antiplatelets are given, gradually causing vessel stenosis [3]. Additional lesions in the dialysis circuit may also have formed over time, resulting in reduced circuit patency [53].

In particular, Björkman et al [44] heavily favoured PTA over DCB in the setting of ACPP. Björkman et al postulated that the younger and more immature AVFs (<1 year compared to the other studies) were more venous than arterial in nature; and with thinner venous walls, local paclitaxel overdose and potential toxicity could be more pronounced, accounting for the poor performance in the DCB arm. The authors stated that the study was discontinued due to slow recruitment; it was also limited by a smaller sample size than necessary for statistical power calculations, slow randomization and possibility of type I error. Insufficient studies provided information on age of AVFs to elucidate an association between age of AVFs and DCB efficacy in our study.

Looking forward, it is critical to determine the optimal concentration of paclitaxel where benefits are best observed. As suggested by the wide intervals between ranking P-Scores, there seems to be some evidence that higher concentrations of paclitaxel may be associated with longer TLPP and ACPP. Of note, among all comparisons against PTA, only DCBs with paclitaxel concentrations of 3.5μg/mm^2^ and 3.0μg/mm^2^ had a statistically significant TLPP advantage. However, upon closer inspection of indirect treatment comparisons generated through the Frequentist network meta-analysis, we did not find a significant difference in TLPP between concentrations of 2.0, 3.0 and 3.5μg/mm^2^.

While specific and potentially influential lesion covariates (such as length, location, type/location of access point and balloon diameter) were not consistently accounted for amongst the RCTs, this may suggest that the true benefit of adding paclitaxel may lie beyond concentrations beyond 3.0μg/mm^2^ – and may be an area of inquiry for further investigations.

### Limitations

This study was not without limitations. In the course of our analyses, we were unable to account for competing risks to restenosis, such as patient mortality, for which previous studies yielded mixed findings. A DCB trial in femoropopliteal stenosis [10] showed increased mortality compared to PTA (24-month fixed-effect RR=1.74, 95%-CI: 1.08-2.81, P=0.02), and a meta-analysis of paclitaxel-coated devices demonstrated an increased mortality by 4.6% at 5-years [51]. That being said, a meta-analysis of paclitaxel-coated devices in dialysis access [54] found no difference in short to midterm mortality among patients compared to PTA (RR=1.26, 95%-CI: 0.85-1.89, P=0.25).

Additionally, although IPD was recovered graphically, the reconstruction algorithm is unable to retrieve patient-level covariate information, which may offer beneficial insight if adjusted for in models utilised. Potentially prognostic covariates for restenosis include vascular access age (as mentioned above), type of access point (AVF or AVG – where the absence of vascular smooth muscle in AVGs may prevent paclitaxel from exerting its desired action), diabetes mellitus [55], lesion length [56], blood flow volume [57], location of the AVF or AVG (forearm versus arm) [58], and type or duration of antiplatelet treatment [59]. Unfortunately, not all studies reported these parameters; there was hence inadequate data for us to perform a meta-regression.

Finally, clinical judgement to proceed with either DCB or PTA should be considered alongside the side effects of paclitaxel, procedure-related morbidity and cost-benefit analysis which are beyond the scope of this forum.

## Conclusion

Hereto, we present the first patient-level, RCT-only meta-analysis comparing DCB to PTA in hemodialysis access restenosis. Pooled-analyses of 11 RCTs comprising 1,243 patients – including 2 recent RCTs published in 2020 – demonstrate that the overall evidence suggests that DCB is favoured over PTA in prolonging TLPP and ACPP.

Intriguingly, we also found some evidence that higher paclitaxel concentrations may be associated with longer TLPP. Future investigations may benefit from exploring more dose-efficacy relationships and may benefit from adjusting for more lesion specific covariates such as age, length, location, type and balloon diameter.

## Supporting information

Supplementary Comparisons 1

Supplementary Comparisons 2

## Data Availability

This manuscript makes use of publicly-available data from published studies, therefore no original or additional data is available for sharing.

## DISCLOSURE

### Contributions

Study design: C.W.T., J.J.Z., K.Y.F.

Data collection: K.Y.F., J.J.Z., E.T.

Statistical analysis: J.J.Z., R.S., N.S., E.T.

Manuscript writing: K.Y.F., J.J.Z.

Critical revision of manuscript: all authors.

Study supervision: C.W.T.

### Ethics

Not applicable.

## PRISMA-IPD Checklist of items to include when reporting a systematic review and meta-analysis of individual participant data (IPD)

| PRISMA-IPD Section/topic | Item No | Checklist item | Reported on page |
| --- | --- | --- | --- |
| **Title** |  |  |  |
| Title | 1 | Identify the report as a systematic review and meta-analysis of individual participant data. | 1 |
| **Abstract** |  |  |  |
| Structured summary | 2 | Provide a structured summary including as applicable:<br>**Background:** state research question and main objectives, with information on participants, interventions, comparators and outcomes.<br>**Methods:** report eligibility criteria; data sources including dates of last bibliographic search or elicitation, noting that IPD were sought; methods of assessing risk of bias.<br>**Results:** provide number and type of studies and participants identified and number (%) obtained; summary effect estimates for main outcomes (benefits and harms) with confidence intervals and measures of statistical heterogeneity. Describe the direction and size of summary effects in terms meaningful to those who would put findings into practice.<br>**Discussion:** state main strengths and limitations of the evidence, general interpretation of the results and any important implications.<br>**Other:** report primary funding source, registration number and registry name for the systematic review and IPD meta-analysis. | 3-4 |
| **Introduction** |  |  |  |
| Rationale | 3 | Describe the rationale for the review in the context of what is already known. | 5 |
| Objectives | 4 | Provide an explicit statement of the questions being addressed with reference, as applicable, to participants, interventions, comparisons, outcomes and study design (PICOS). Include any hypotheses that relate to particular types of participant-level subgroups. | 5 |
| **Methods** |  |  |  |
| Protocol and registration | 5 | Indicate if a protocol exists and where it can be accessed. If available, provide registration information including registration number and registry name. Provide publication details, if applicable. | NA |
| Eligibility criteria | 6 | Specify inclusion and exclusion criteria including those relating to participants, interventions, comparisons, outcomes, study design and characteristics (e.g. years when conducted, required minimum follow-up). Note whether these were applied at the study or individual level i.e. whether eligible participants were included (and ineligible participants excluded) from a study that included a wider population than specified by the review inclusion criteria. The rationale for criteria should be stated. | 6 |
| Identifying studies - | 7 | Describe all methods of identifying published and unpublished studies including, as applicable: which bibliographic databases were searched with dates of coverage; details of any hand searching including of conference proceedings; use of study registers | 6 |
| information sources |  | and agency or company databases; contact with the original research team and experts in the field; open adverts and surveys. Give the date of last search or elicitation. |  |
| Identifying studies - search | 8 | Present the full electronic search strategy for at least one database, including any limits used, such that it could be repeated. | 6 |
| Study selection processes | 9 | State the process for determining which studies were eligible for inclusion. | 6 |
| Data collection processes | 10 | Describe how IPD were requested, collected and managed, including any processes for querying and confirming data with investigators. If IPD were not sought from any eligible study, the reason for this should be stated (for each such study). | 6-8 |
|  |  | If applicable, describe how any studies for which IPD were not available were dealt with. This should include whether, how and what aggregate data were sought or extracted from study reports and publications (such as extracting data independently in duplicate) and any processes for obtaining and confirming these data with investigators. |  |
| Data items | 11 | Describe how the information and variables to be collected were chosen. List and define all study level and participant level data that were sought, including baseline and follow-up information. If applicable, describe methods of standardising or translating variables within the IPD datasets to ensure common scales or measurements across studies. | 6-8 |
| IPD integrity | A1 | Describe what aspects of IPD were subject to data checking (such as sequence generation, data consistency and completeness, baseline imbalance) and how this was done. | 6-8 |
| Risk of bias assessment in individual studies. | 12 | Describe methods used to assess risk of bias in the individual studies and whether this was applied separately for each outcome. If applicable, describe how findings of IPD checking were used to inform the assessment. Report if and how risk of bias assessment was used in any data synthesis. | 6 |
| Specification of outcomes and effect measures | 13 | State all treatment comparisons of interests. State all outcomes addressed and define them in detail. State whether they were pre-specified for the review and, if applicable, whether they were primary/main or secondary/additional outcomes. Give the principal measures of effect (such as risk ratio, hazard ratio, difference in means) used for each outcome. | 6-8 |
| Synthesis methods | 14 | Describe the meta-analysis methods used to synthesise IPD. Specify any statistical methods and models used. Issues should include (but are not restricted to):<br>• Use of a one-stage or two-stage approach.<br>• How effect estimates were generated separately within each study and combined across studies (where applicable).<br>• Specification of one-stage models (where applicable) including how clustering of patients within studies was accounted for.<br>• Use of fixed or random effects models and any other model assumptions, such as proportional hazards.<br>• How (summary) survival curves were generated (where applicable).<br>• Methods for quantifying statistical heterogeneity (such as $I^2$ and $\tau^2$). | 6-8 |
|  |  | How studies providing IPD and not providing IPD were analysed together (where applicable).<br>How missing data within the IPD were dealt with (where applicable). |  |
| Exploration of variation in effects | A2 | If applicable, describe any methods used to explore variation in effects by study or participant level characteristics (such as estimation of interactions between effect and covariates). State all participant-level characteristics that were analysed as potential effect modifiers, and whether these were pre-specified. | 6-8 |
| Risk of bias across studies | 15 | Specify any assessment of risk of bias relating to the accumulated body of evidence, including any pertaining to not obtaining IPD for particular studies, outcomes or other variables. | 6 |
| Additional analyses | 16 | Describe methods of any additional analyses, including sensitivity analyses. State which of these were pre-specified. | 6-8 |
| **Results** |  |  |  |
| Study selection and IPD obtained | 17 | Give numbers of studies screened, assessed for eligibility, and included in the systematic review with reasons for exclusions at each stage. Indicate the number of studies and participants for which IPD were sought and for which IPD were obtained. For those studies where IPD were not available, give the numbers of studies and participants for which aggregate data were available. Report reasons for non-availability of IPD. Include a flow diagram. | 9 |
| Study characteristics | 18 | For each study, present information on key study and participant characteristics (such as description of interventions, numbers of participants, demographic data, unavailability of outcomes, funding source, and if applicable duration of follow-up). Provide (main) citations for each study. Where applicable, also report similar study characteristics for any studies not providing IPD. | 9 |
| IPD integrity | A3 | Report any important issues identified in checking IPD or state that there were none. | 9 |
| Risk of bias within studies | 19 | Present data on risk of bias assessments. If applicable, describe whether data checking led to the up-weighting or down-weighting of these assessments. Consider how any potential bias impacts on the robustness of meta-analysis conclusions. | 9 |
| Results of individual studies | 20 | For each comparison and for each main outcome (benefit or harm), for each individual study report the number of eligible participants for which data were obtained and show simple summary data for each intervention group (including, where applicable, the number of events), effect estimates and confidence intervals. These may be tabulated or included on a forest plot. | Table 1A, 1B |
| Results of syntheses | 21 | Present summary effects for each meta-analysis undertaken, including confidence intervals and measures of statistical heterogeneity. State whether the analysis was pre-specified, and report the numbers of studies and participants and, where applicable, the number of events on which it is based. | Figure 2, 9-11 |
|  |  | When exploring variation in effects due to patient or study characteristics, present summary interaction estimates for each characteristic examined, including confidence intervals and measures of statistical heterogeneity. State whether the analysis was pre-specified. State whether any interaction is consistent across trials. |  |
|  |  | Provide a description of the direction and size of effect in terms meaningful to those who would put findings into practice. |  |
| Risk of bias across studies | 22 | Present results of any assessment of risk of bias relating to the accumulated body of evidence, including any pertaining to the availability and representativeness of available studies, outcomes or other variables. | Table 2B, 9 |
| Additional analyses | 23 | Give results of any additional analyses (e.g. sensitivity analyses). If applicable, this should also include any analyses that incorporate aggregate data for studies that do not have IPD. If applicable, summarise the main meta-analysis results following the inclusion or exclusion of studies for which IPD were not available. | Figure 3, 4A-E, 5A-B |
| **Discussion** |  |  |  |
| Summary of evidence | 24 | Summarise the main findings, including the strength of evidence for each main outcome. | 9-11 |
| Strengths and limitations | 25 | Discuss any important strengths and limitations of the evidence including the benefits of access to IPD and any limitations arising from IPD that were not available. | 13-14 |
| Conclusions | 26 | Provide a general interpretation of the findings in the context of other evidence. | 14 |
| Implications | A4 | Consider relevance to key groups (such as policy makers, service providers and service users). Consider implications for future research. | 13-14 |
| **Funding** |  |  |  |
| Funding | 27 | Describe sources of funding and other support (such as supply of IPD), and the role in the systematic review of those providing such support. | NA |

**A1 – A3 denote new items that are additional to standard PRISMA items. A4 has been created as a result of re-arranging content of the standard PRISMA statement to suit the way that systematic review IPD meta-analyses are reported.**
© Reproduced with permission of the PRISMA IPD Group, which encourages sharing and reuse for non-commercial purposes

## PRISMA NMA Checklist of Items to Include When Reporting A Systematic Review Involving a Network Meta-analysis

| Section/Topic | Item # | Checklist Item | Reported on Page # |
| --- | --- | --- | --- |
| **TITLE** |  |  |  |
| Title | 1 | Identify the report as a systematic review *incorporating a network meta-analysis (or related form of meta-analysis)*. | 1 |
| **ABSTRACT** |  |  |  |
| Structured summary | 2 | Provide a structured summary including, as applicable:<br>**Background:** main objectives<br>**Methods:** data sources; study eligibility criteria, participants, and interventions; study appraisal; and *synthesis methods, such as network meta-analysis*.<br>**Results:** number of studies and participants identified; summary estimates with corresponding confidence/credible intervals; *treatment rankings may also be discussed. Authors may choose to summarize pairwise comparisons against a chosen treatment included in their analyses for brevity.*<br>**Discussion/Conclusions:** limitations; conclusions and implications of findings.<br>**Other:** primary source of funding; systematic review registration number with registry name. | 3-4 |
| **INTRODUCTION** |  |  |  |
| Rationale | 3 | Describe the rationale for the review in the context of what is already known, *including mention of why a network meta-analysis has been conducted*. | 5 |
| Objectives | 4 | Provide an explicit statement of questions being addressed, with reference to participants, interventions, comparisons, outcomes, and study design (PICOS). | 5 |
| **METHODS** |  |  |  |
| Protocol and registration | 5 | Indicate whether a review protocol exists and if and where it can be accessed (e.g., Web address); and, if available, provide registration information, including registration number. | NA |
| Eligibility criteria | 6 | Specify study characteristics (e.g., PICOS, length of follow-up) and report characteristics (e.g., years considered, language, publication status) used as criteria for eligibility, giving rationale. *Clearly describe eligible treatments included in the treatment network, and note whether any have been clustered or merged into the same node (with justification).* | 6 |
| Information sources | 7 | Describe all information sources (e.g., databases with dates of coverage, contact with study authors to identify additional studies) in the search and date last searched. | 6 |
| Search | 8 | Present full electronic search strategy for at least one database, including any limits used, such that it could be repeated. | 6 |
| Study selection | 9 | State the process for selecting studies (i.e., screening, eligibility, included in systematic review, and, if applicable, included in the meta-analysis). | 6 |
| Data collection process | 10 | Describe method of data extraction from reports (e.g., piloted forms, independently, in duplicate) and any processes for obtaining and confirming data from investigators. | 6-8 |
| Data items | 11 | List and define all variables for which data were sought (e.g., PICOS, funding sources) and any assumptions and simplifications made. | 6-8 |
| **Geometry of the network** | **S1** | Describe methods used to explore the geometry of the treatment network under study and potential biases related to it. This should include how the evidence base has been graphically summarized for presentation, and what characteristics were compiled and used to describe the evidence base to readers. | 6-8 |
| Risk of bias within individual studies | 12 | Describe methods used for assessing risk of bias of individual studies (including specification of whether this was done at the study or outcome level), and how this information is to be used in any data synthesis. | 6 |
| Summary measures | 13 | State the principal summary measures (e.g., risk ratio, difference in means). *Also describe the use of additional summary measures assessed, such as treatment rankings and surface under the cumulative ranking curve (SUCRA) values, as well as modified approaches used to present summary findings from meta-analyses.* | NA |
| Planned methods of analysis | 14 | Describe the methods of handling data and combining results of studies for each network meta-analysis. This should include, but not be limited to:<br>• *Handling of multi-arm trials;*<br>• *Selection of variance structure;*<br>• *Selection of prior distributions in Bayesian analyses; and*<br>• *Assessment of model fit.* | 6-8 |
| **Assessment of Inconsistency** | **S2** | Describe the statistical methods used to evaluate the agreement of direct and indirect evidence in the treatment network(s) studied. Describe efforts taken to address its presence when found. | 6-8 |
| Risk of bias across studies | 15 | Specify any assessment of risk of bias that may affect the cumulative evidence (e.g., publication bias, selective reporting within studies). | 6 |
| Additional analyses | 16 | Describe methods of additional analyses if done, indicating which were pre-specified. This may include, but not be limited to, the following:<br>• Sensitivity or subgroup analyses;<br>• Meta-regression analyses;<br>• *Alternative formulations of the treatment network; and*<br>• *Use of alternative prior distributions for Bayesian analyses (if applicable).* | 6-8 |
| **RESULTS†** |  |  |  |
| Study selection | 17 | Give numbers of studies screened, assessed for eligibility, and included in the review, with reasons for exclusions at each stage, ideally with a flow diagram. | 9 |
| **Presentation of network structure** | **S3** | Provide a network graph of the included studies to enable visualization of the geometry of the treatment network. | NA |
| **Summary of network geometry** | **S4** | Provide a brief overview of characteristics of the treatment network. This may include commentary on the abundance of trials and randomized patients for the different interventions and pairwise comparisons in the network, gaps of evidence in the treatment network, and potential biases reflected by the network structure. | 9 |
| Study characteristics | 18 | For each study, present characteristics for which data were extracted (e.g., study size, PICOS, follow-up period) and provide the citations. | 9 |
| Risk of bias within studies | 19 | Present data on risk of bias of each study and, if available, any outcome level assessment. | 9 |
| Results of individual studies | 20 | For all outcomes considered (benefits or harms), present, for each study: 1) simple summary data for each intervention group, and 2) effect estimates and confidence intervals. *Modified approaches may be needed to deal with information from larger networks.* | Table 1A, 1B |
| Synthesis of results | 21 | Present results of each meta-analysis done, including confidence/credible intervals. *In larger networks, authors may focus on comparisons versus a particular comparator (e.g. placebo or standard care), with full findings presented in an appendix. League tables and forest plots may be considered to summarize pairwise comparisons.* If additional summary measures were explored (such as treatment rankings), these should also be presented. | 9-11 |
| **Exploration for inconsistency** | **S5** | Describe results from investigations of inconsistency. This may include such information as measures of model fit to compare consistency and inconsistency models, *P* values from statistical tests, or summary of inconsistency estimates from different parts of the treatment network. | 10-11 |
| Risk of bias across studies | 22 | Present results of any assessment of risk of bias across studies for the evidence base being studied. | Table 2B, 9 |
| Results of additional analyses | 23 | Give results of additional analyses, if done (e.g., sensitivity or subgroup analyses, meta-regression analyses, *alternative network geometries studied, alternative choice of prior distributions for Bayesian analyses*, and so forth). | Figure 3, 4A-E, 5A-B |
| **DISCUSSION** |  |  |  |
| Summary of evidence | 24 | Summarize the main findings, including the strength of evidence for each main outcome; consider their relevance to key groups (e.g., healthcare providers, users, and policy-makers). | 12-13 |
| Limitations | 25 | Discuss limitations at study and outcome level (e.g., risk of bias), and at review level (e.g., incomplete retrieval of identified research, reporting bias). *Comment on the validity of the assumptions, such as transitivity and consistency. Comment on any concerns regarding network geometry (e.g., avoidance of certain comparisons).* | 13-14 |
| Conclusions | 26 | Provide a general interpretation of the results in the context of other evidence, and implications for future research. | 14 |
| **FUNDING** |  |  | NA |
| Funding | 27 | Describe sources of funding for the systematic review and other support (e.g., supply of data); role of funders for the systematic review. This should also include information regarding whether funding has been received from manufacturers of treatments in the network and/or whether some of the authors are content experts with professional conflicts of interest that could affect use of treatments in the network. |  |

PICOS = population, intervention, comparators, outcomes, study design.
\* Text in italics indicates wording specific to reporting of network meta-analyses that has been added to guidance from the PRISMA statement.
† Authors may wish to plan for use of appendices to present all relevant information in full detail for items in this section.

