## Supplementary Materials for "Drug-coated balloons for dysfunctional hemodialysis venous access: A patient-level meta-analysis of randomized controlled trials"

TLPP, target lesion primary patency

ACPP, access circuit primary patency

DCB, drug coated balloon

PTA, percutaneous transluminal angioplasty

HR, hazard ratio

95CI, 95% confidence interval

GT test, Grambsch-Therneau test

NR, not reported

RMST, restricted mean survival time

### Supplementary Table 1 Electronic literature search algorithm

(‘Arteriovenous Graft’ OR ‘Arteriovenous Fistula’ OR ‘Arteriovenous Shunt’ OR ‘Vascular access’ OR ‘Dialysis access’ OR ‘Arteriovenous Anastomosis’ OR ‘Arteriovenous Shunt’ OR ‘Blood Vessel Prosthesis’) AND (‘Drug eluting balloon’ OR 'Drug-eluting balloon' OR ‘Paclitaxel coated balloon’ OR 'Paclitaxel-coated balloon' OR ‘Drug coated balloon’ OR 'Drug-coated balloon' OR ‘Paclitaxel’) AND ('Random' OR 'Randomized' OR 'Randomised')”

### Supplementary Table 2 Comparisons to original values

**A - Target lesion primary patency (TLPP)**

| **Ref** | **HR** | **P-value** | **GT test** | **Study reported HR** | **Study reported P-value** |
| --- | --- | --- | --- | --- | --- |
| **Irani, 2018** | 1.621467 | 0.045813 | 0.907658 | 1.626 | 0.047 |
| **Kim, 2020** | 1.074071 | 0.88035 | 0.533885 | NR | NR |
| **Kitrou, 2015-A** | 1.921684 | 0.062431 | 0.6078 | 2.092 | 0.03 |
| **Kitrou, 2015-B** | 0.295983 | 0.000865 | 0.528433 | 0.23 | 0.0007 |
| **Liao, 2019** | 2.14591813 | 0.02043321 | 0.26538794 | 2.096 | 0.019 |
| **Lookstein, 2020** | 0.371298 | 9.59E-06 | 0.970837 | 0.39 | NR |
| **Morenosanchez, 2020** | 1.216337 | 0.426318 | 0.402269 | NR | NR |
| **Roosen, 2017** | 0.599314 | 0.168136 | 0.672421 | NR | NR |
| **Swinnen, 2018** | 2.52898344 | 0.00079699 | 0.11610912 | 2.44 | 0.002 |
| **Trerotola, 2019** | 1.1984307 | 0.21949194 | 0.09874912 | NR | NR |

**B - Access circuit primary patency (ACPP)**

| **Ref** | **HR** | **P-value** | **GT test** | **Study reported HR** | **Study reported P-value** |
| --- | --- | --- | --- | --- | --- |
| **Bjorkman, 2018** | 0.24527633 | 0.00063836 | 0.22290895 | NR | NR |
| **Irani, 2018** | 1.42449675 | 0.12904446 | 0.91164011 | 1.431 | 0.13 |
| **Kitrou, 2015-A** | 1.99763573 | 0.04514467 | 0.92855361 | 2.088 | 0.04 |
| **Liao, 2019** | 0.50380321 | 0.03474443 | 0.12043909 | 0.55187 | 0.056 |
| **Lookstein, 2020** | 2.34339732 | 8.26E-06 | 0.18956994 | NR | NR |
| **Trerotola, 2019** | 1.1169837 | 0.42867047 | 0.82308445 | NR | NR |

Some HR values may be inverted for ease of reference.

### Supplementary Table 3 Joanna Briggs Institute critical appraisal tool for randomized controlled trials

|  | Kitrou-A, 2015 | Kitrou-B, 2015 | Roosen, 2017 | Trerotola, 2019 | Björkman, 2018 | Swinnen, 2018 | Irani, 2018 | Liao, 2019 | Moreno-Sanchez, 2020 | Lookstein, 2020 | Kim, 2020 |
| --- | --- | --- | --- | --- | --- | --- | --- | --- | --- | --- | --- |
| Was true randomization used for assignment of participants to treatment groups? | Y | Y | Y | Y | Y | Y | Y | Y | Y | Y | Y |
| Was allocation to treatment groups concealed? | Y | Y | Y | Y | Y | Y | Y | Y | Y | Y | Y |
| Were treatment groups similar at the baseline?^ | Y | Y | N (1) | Y | Y | Y | N (1) | N (2) | N (1) | Y | Y |
| Were participants blind to treatment assignment? | N | Y | Y | Y | Y | Y | N | Y | Y | Y | Y |
| Were those delivering treatment blind to treatment assignment? | N | N | N | N | N | N | N | N | N | N | N |
| Were outcomes assessors blind to treatment assignment? | N | N | Y | Y | Y | Y | N | Y | N | N | N |
| Were treatment groups treated identically other than the intervention of interest? | Y | Y | Y | Y | Y | Y | Y | Y | Y | Y | Y |
| Was follow up complete and if not, were differences between groups in terms of their follow up adequately described and analysed? | Y | Y | Y | N, Y | Y | N, Y | N, Y | Y | N, Y | N, Y | N, Y |
| Were participants analysed in the groups to which they were randomized? | Y | Y | Y | NA* | NA* | NA* | NA* | Y | NA* | NA* | Y |
| Were outcomes measured in the same way for treatment groups? | Y | Y | Y | Y | Y | Y | Y | Y | Y | Y | Y |
| Were outcomes measured in a reliable way? | Y | Y | Y | Y | Y | Y | Y | Y | Y | Y | Y |
| Was appropriate statistical analysis used? | Y | Y | Y | Y | N (lacked statistical power) | Y | Y | Y | Y | Y | Y |
| Was the trial design appropriate, and any deviations from the standard randomized controlled trial design (individual randomization, parallel groups) accounted for in the conduct and analysis of the trial? | Y | Y | Y | Y | N (study halted after only 39 patients) | Y | Y | Y | Y | Y | Y |

^Numbers in parentheses show the number of baseline parameters listed in the article, if any, that differed between the experimental and control groups.

* For trials that did per-protocol analysis instead of intention-to-treat analysis, we rated them as ‘NA’ for this question.

### Supplementary Figure 1 Schoenfeld Residuals

| **A - TLPP** | **B - ACPP** |
| --- | --- |
| 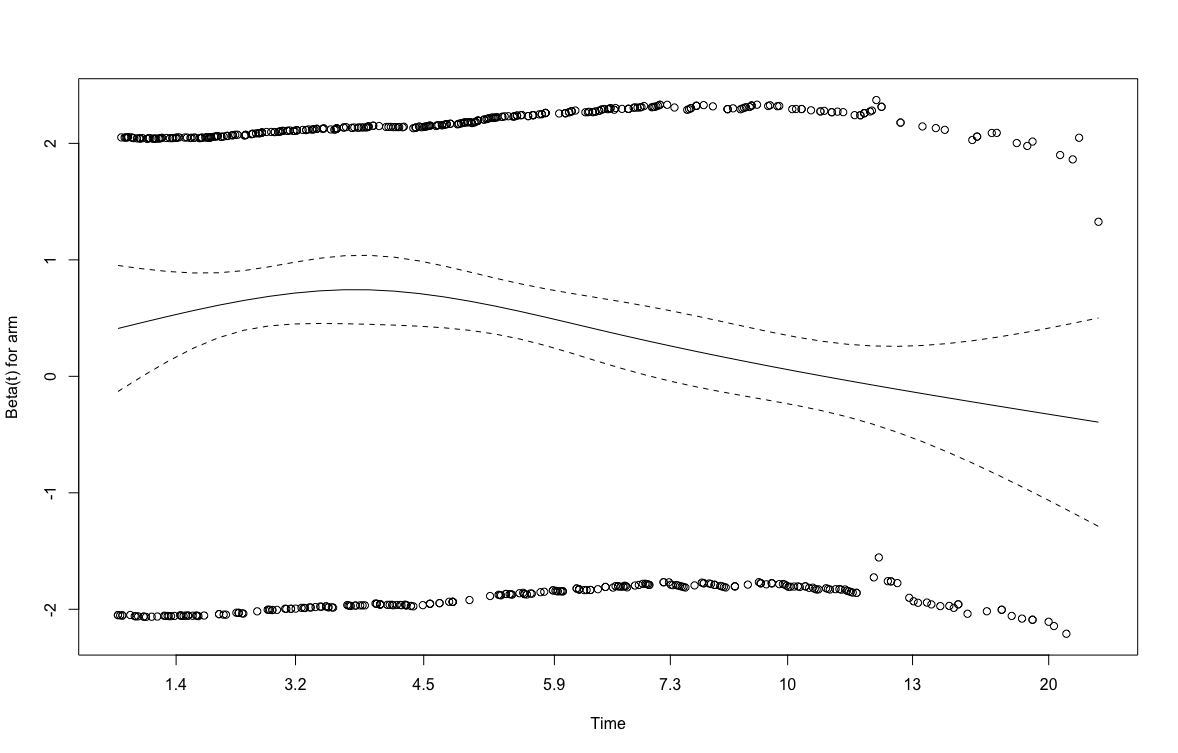  GT test, P = 0.0053 | 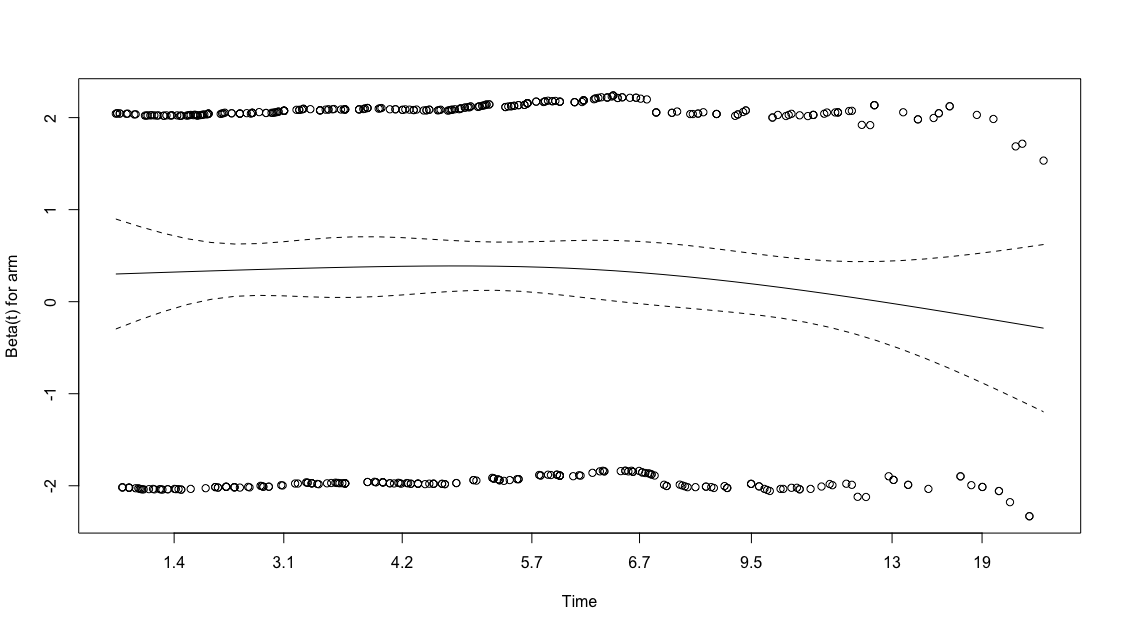  GT test, P = 0.300 |

### Supplementary Figure 2 RMST

Restricted Mean Survival Time for Target Lesion Primary Patency at (1-, 2-, 3- years)

| **A - 1 year** | **B - 2 year** |
| --- | --- |
| 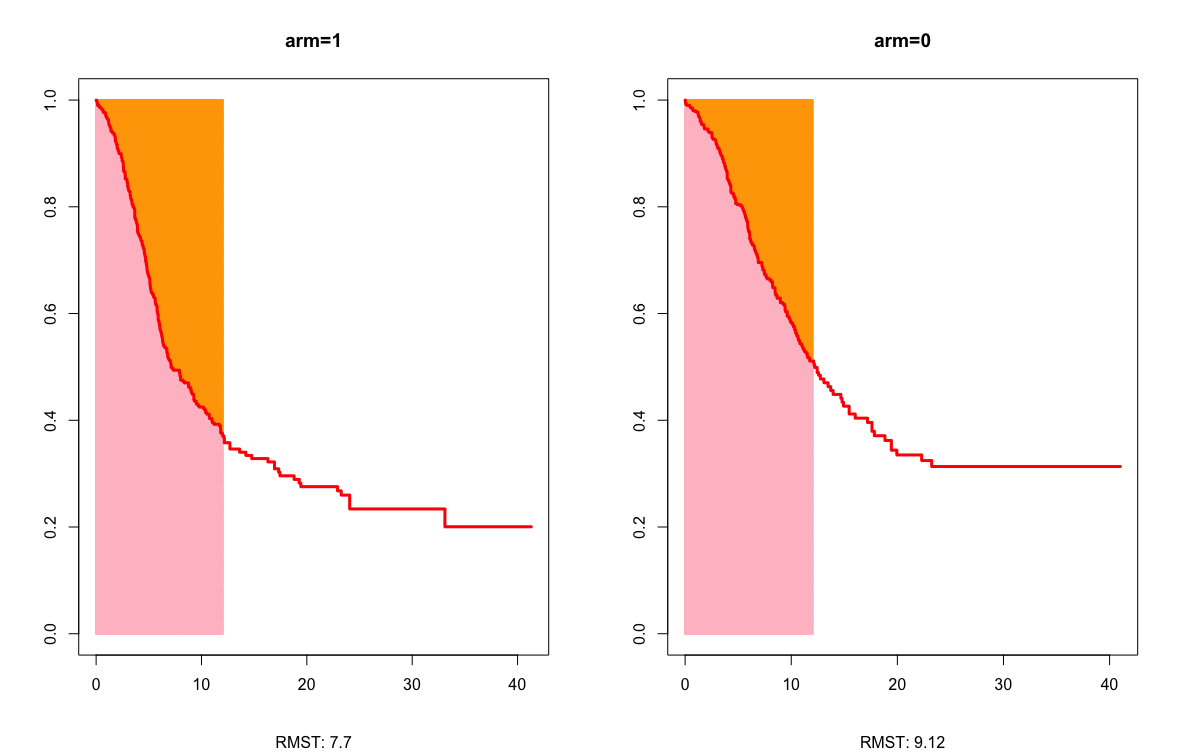 | 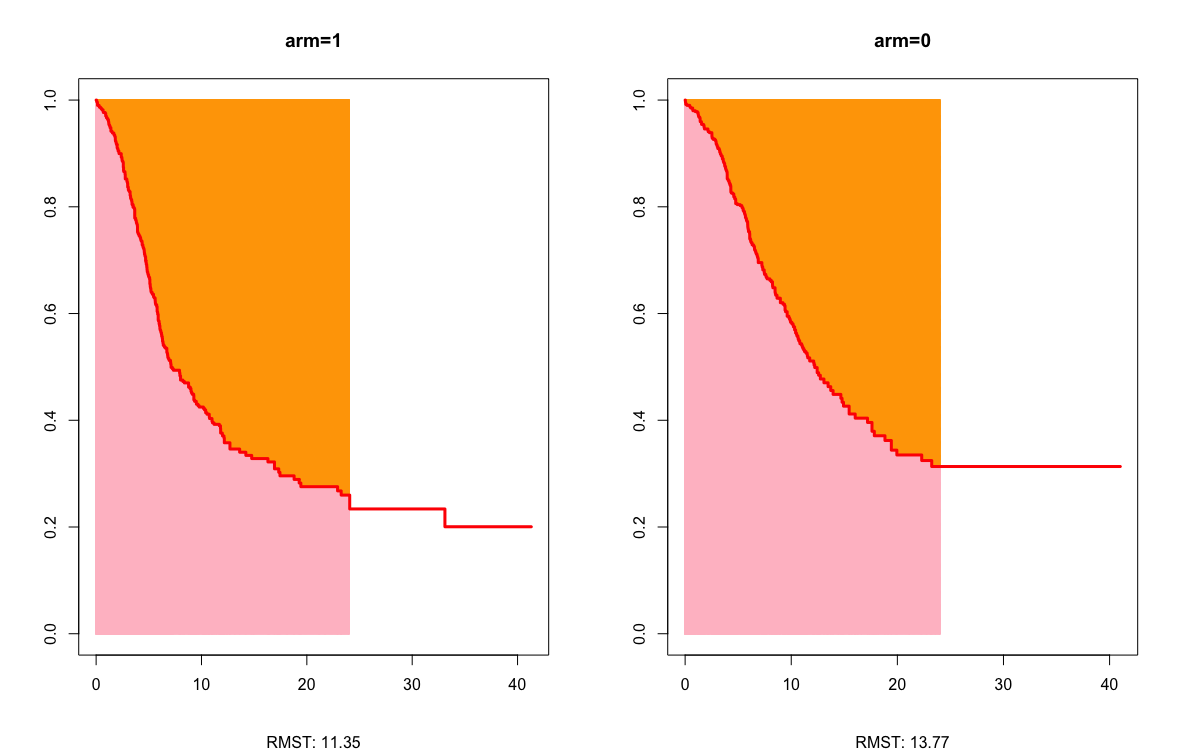 |
| **C - 3 year** |  |
| 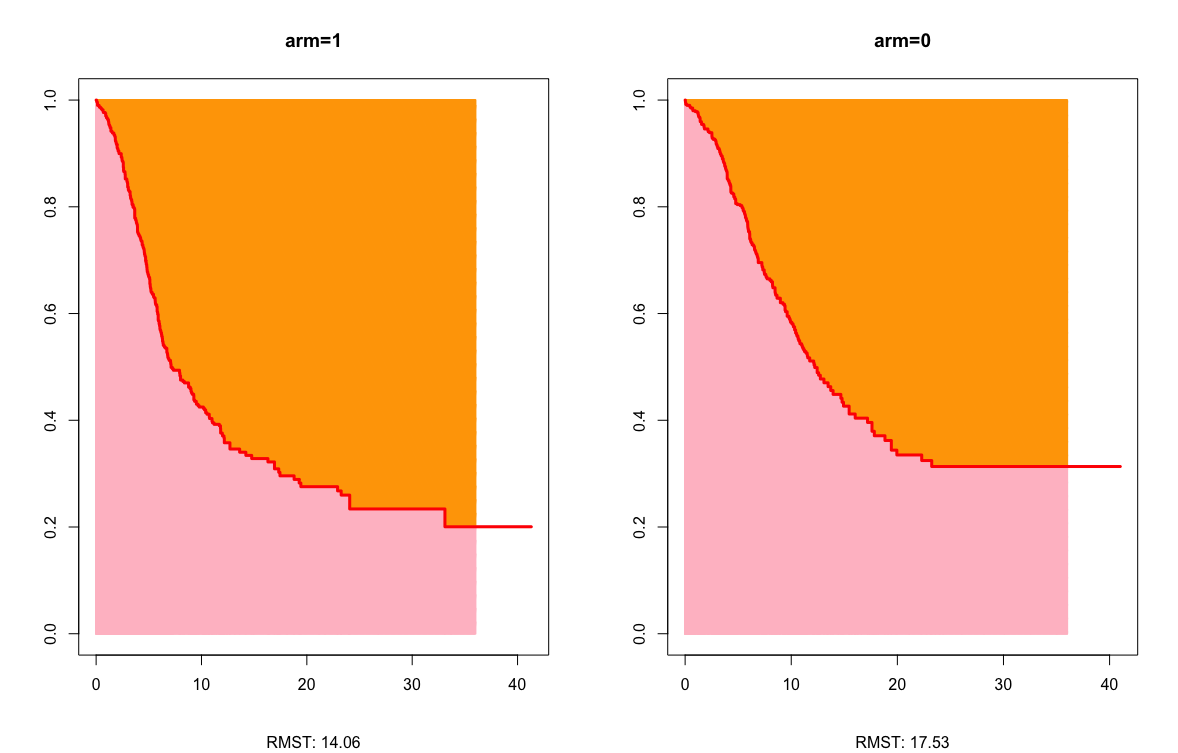 |  |

Arm=0 refers to DCB; Arm=1 refers to PTA.

### Supplementary Figure 3 Cure Models

Cure models

| **A - PTA (TLPP)** | **B - DCB (TLPP)** |
| --- | --- |
| 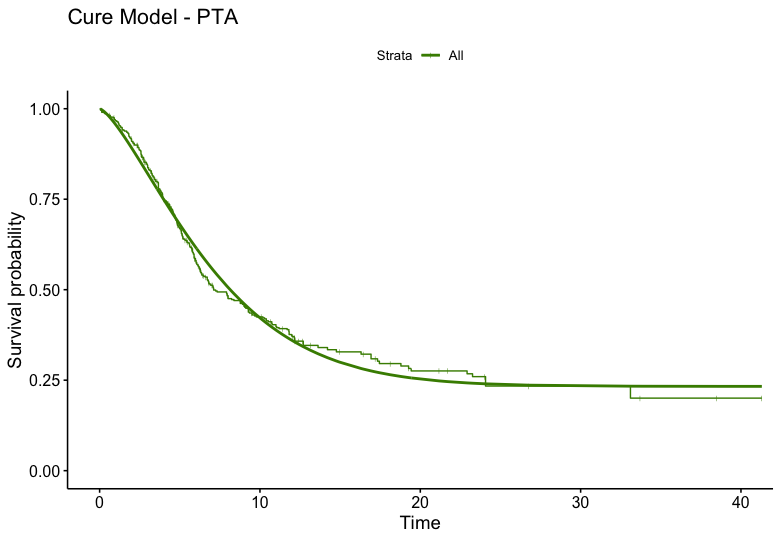 | 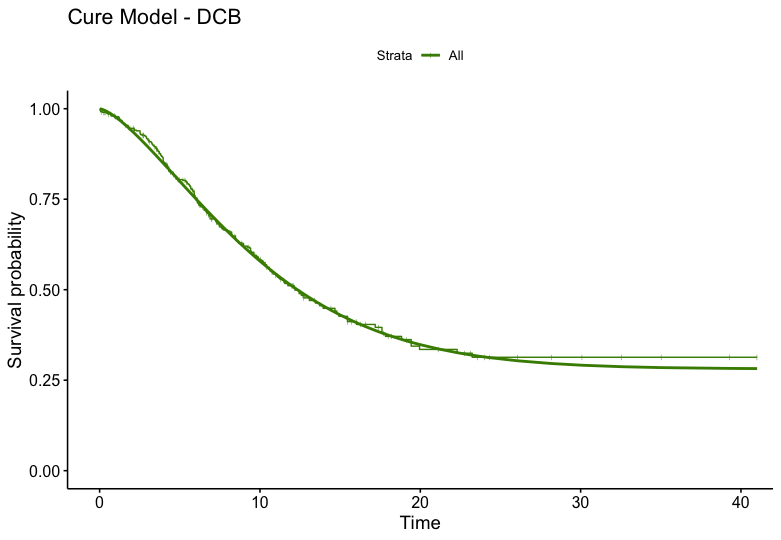 |

### Supplementary Figure 4 Funnel plots for conventional two-stage Frequentist Meta-Analysis

| **A - TLPP** | **B - ACPP** |
| --- | --- |
| 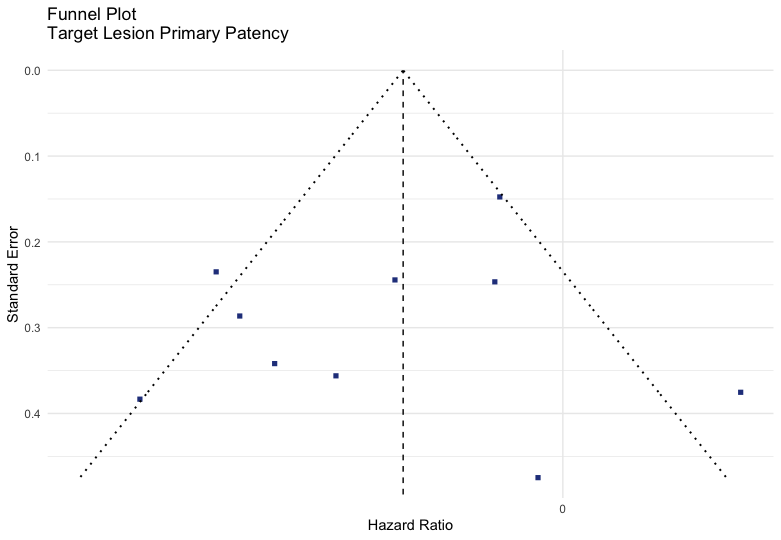  Egger’s Test = 0.507 | 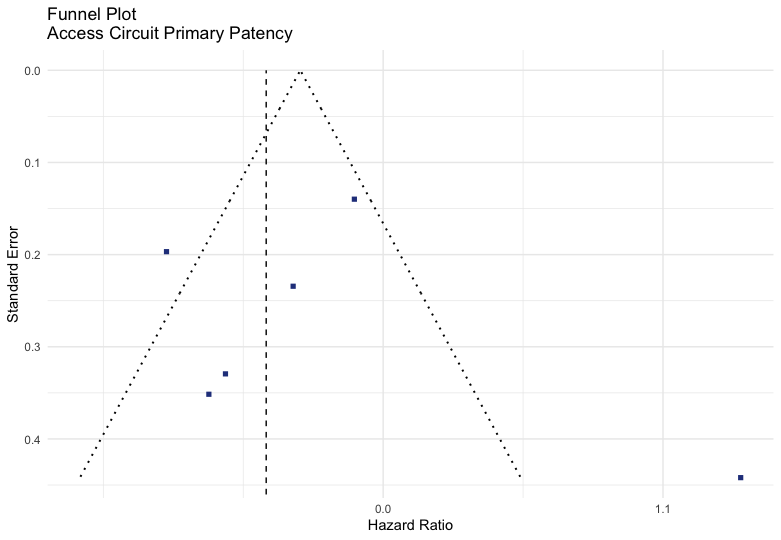  Egger’s Test = 0.771 |

### Supplementary Figure 5A Baujat plot

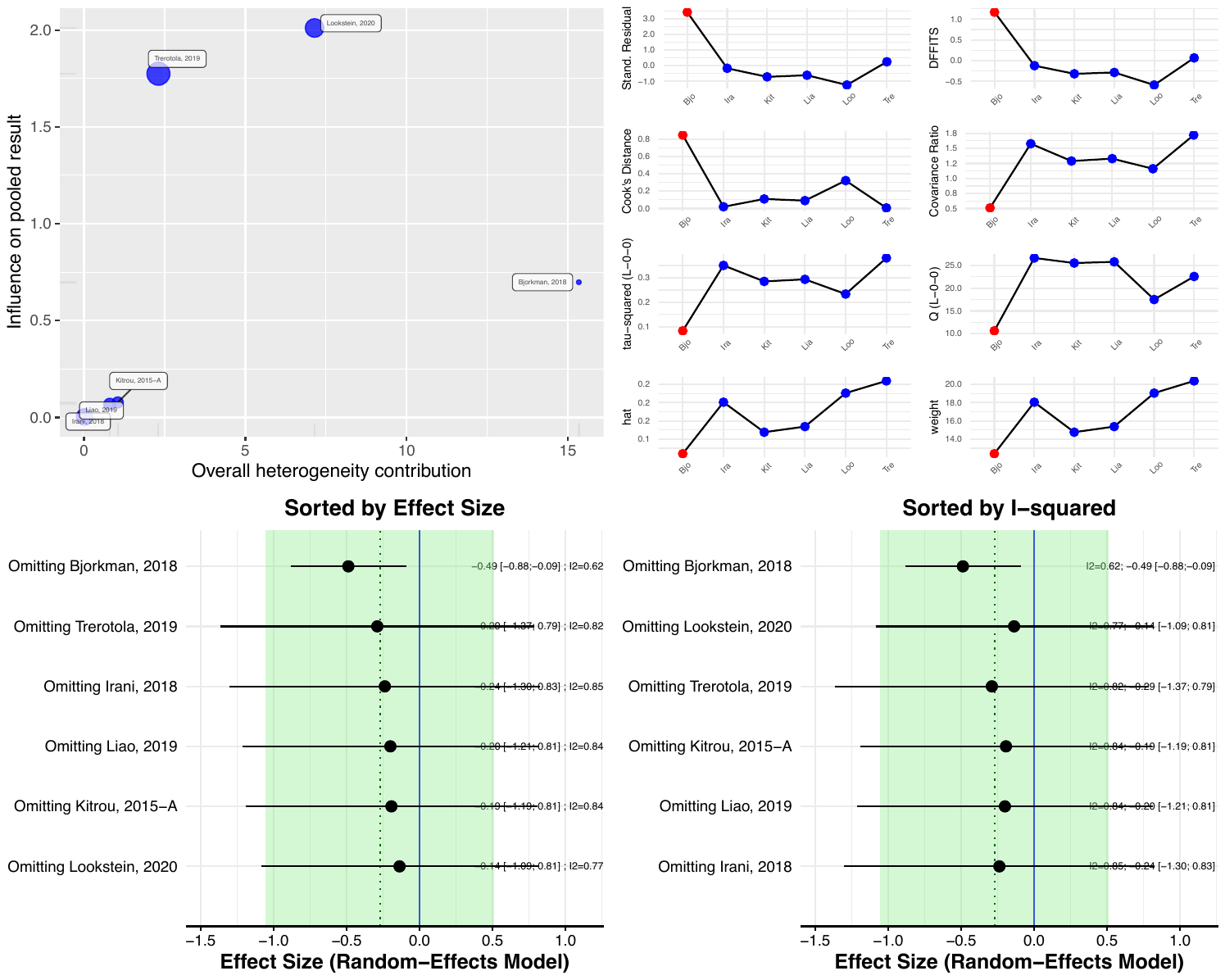

### Supplementary Figure 5B Influence analysis

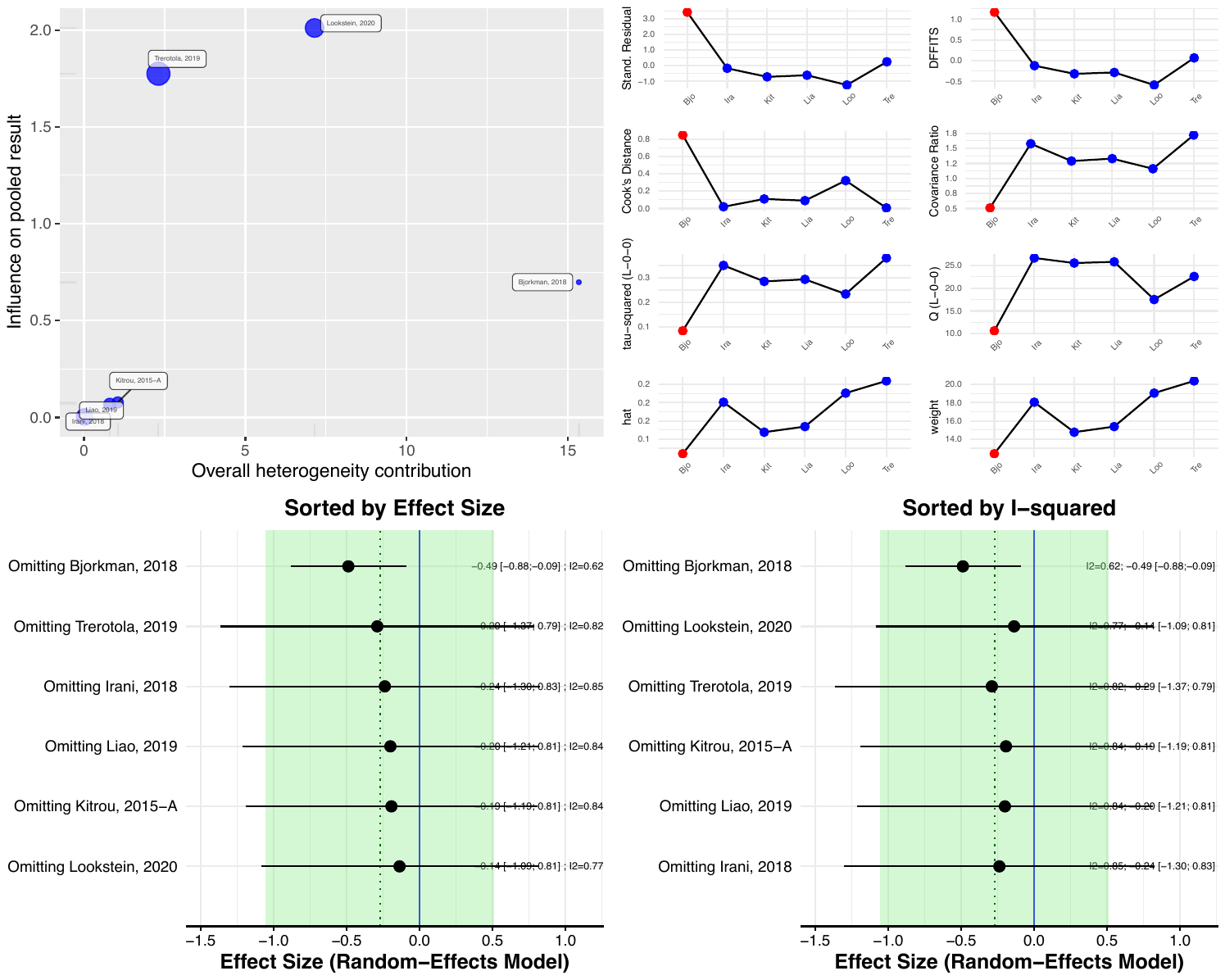

dffits: The DFFITS value of a study indicates in standard deviations how much the predicted pooled effect changes after excluding this study.

cook.d: Cook’s distance resembles the Mahalanobis distance which may be interpreted as the distance between the value once the study is included compared to when it is excluded.

cov.r: The covariance ratio is the determinant of the variance-covariance matrix of the parameter estimates when the study is removed, divided by the determinant of the variance-covariance matrix of the parameter estimates when the full dataset is considered.

### Supplementary Figure 5C Leave-one-out analysis

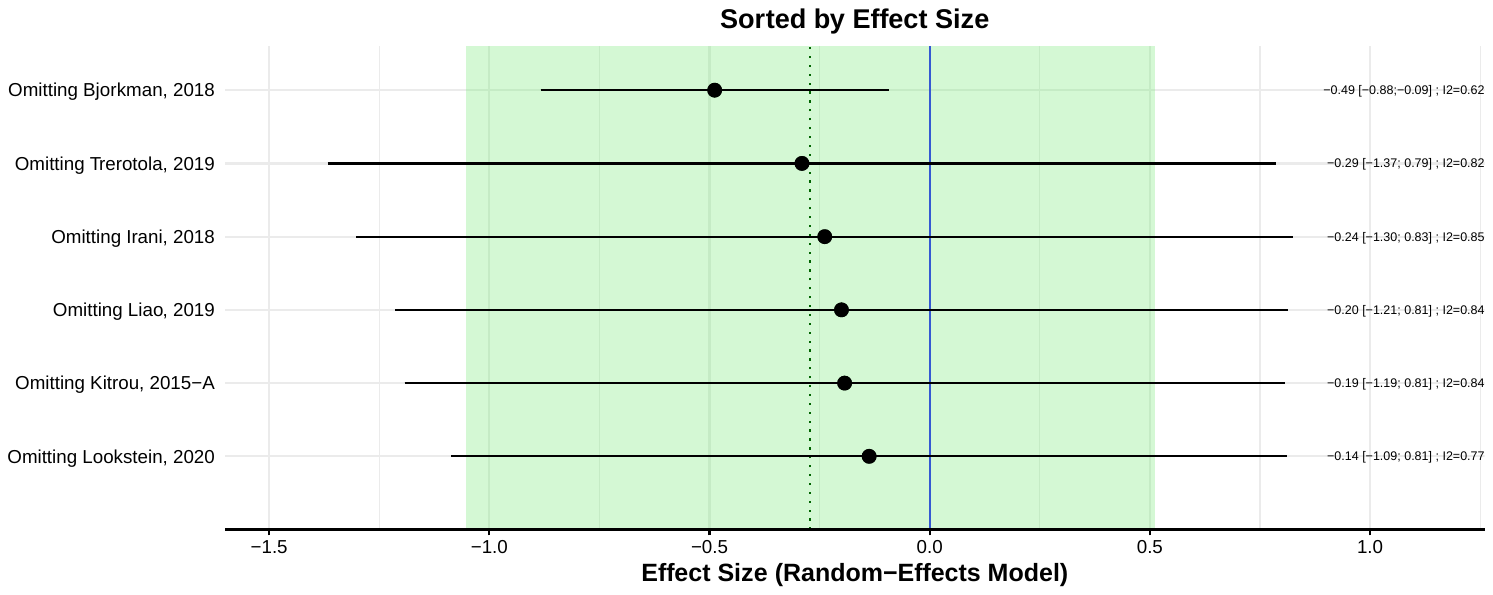

### Supplementary Figure 6 Subgroup analysis within one-stage meta-analysis

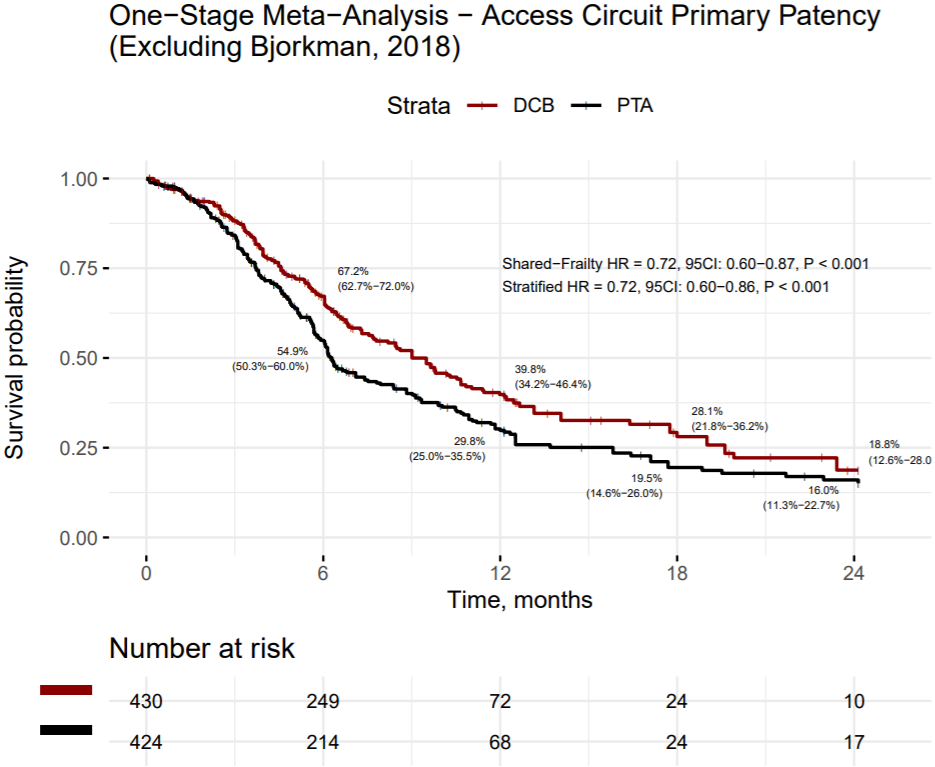

### Supplementary Figure 7 Funnel plots for Network Meta-Analysis

| **A - TLPP** | **B - ACPP** |
| --- | --- |
| 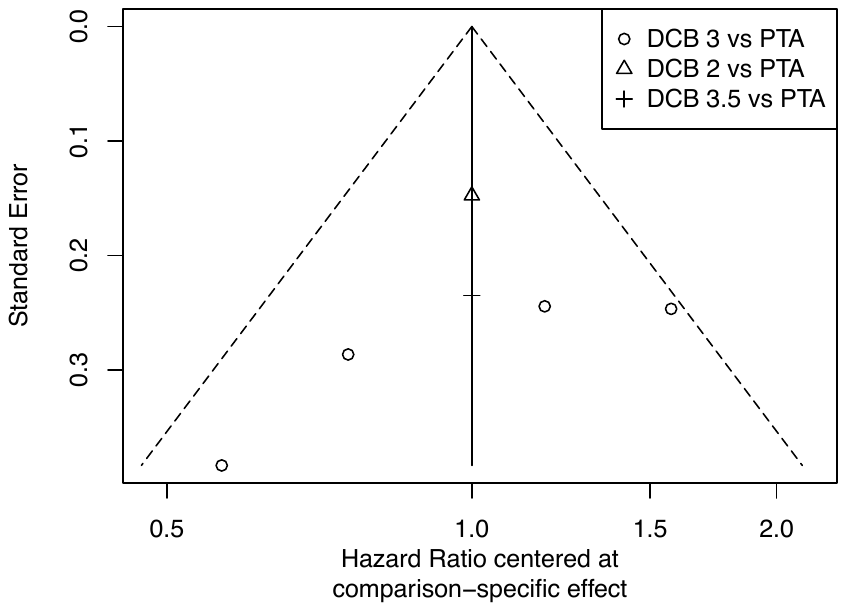 | 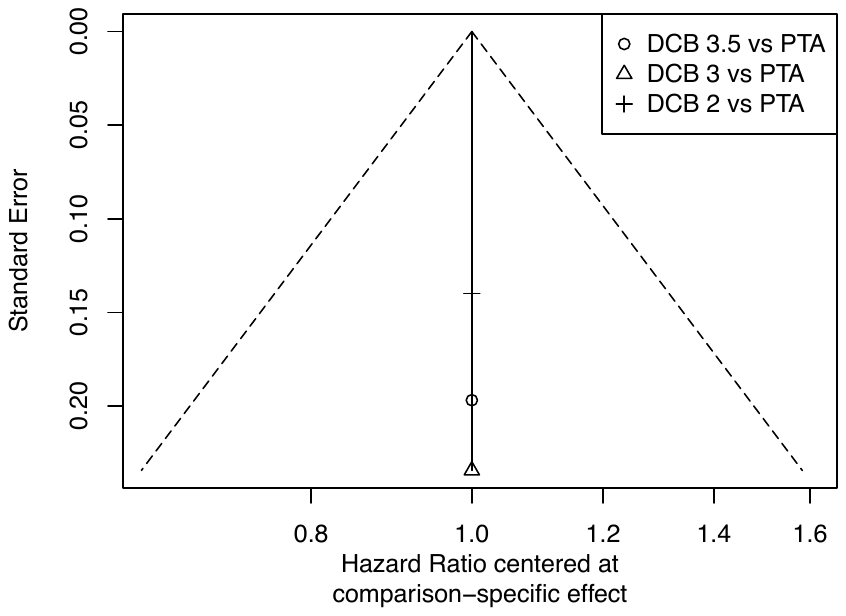 |

The numerical value refers to the concentration of paclitaxel used, for example, DCB 2 = drug coated balloon with 2.0 μg/mm^2^ of paclitaxel.

### Supplementary Figure 8 Network plots of Network Meta-Analysis

| **A - TLPP** | **B - ACPP** |
| --- | --- |
| 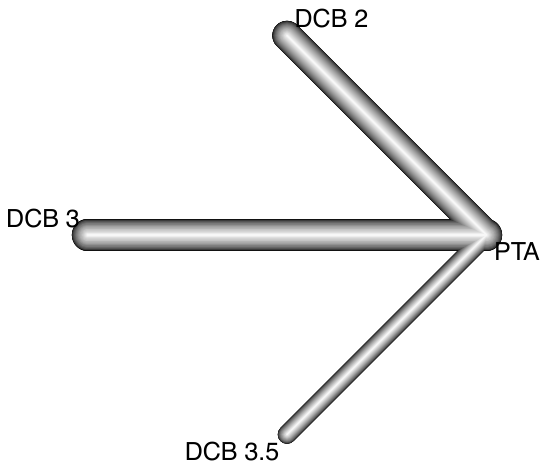 | 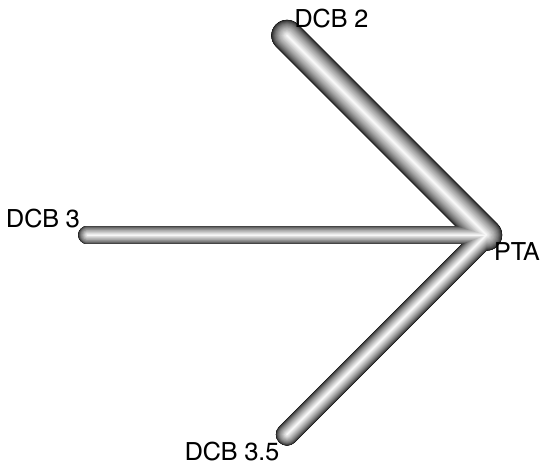 |

The numerical value refers to the concentration of paclitaxel used, for example, DCB 2 = drug coated balloon with 2.0 μg/mm^2^ of paclitaxel.
