## Supplementary Comparisons 1 for "Drug-coated balloons for dysfunctional hemodialysis venous access: A patient-level meta-analysis of randomized controlled trials"

### Irani (TLPP), 2018

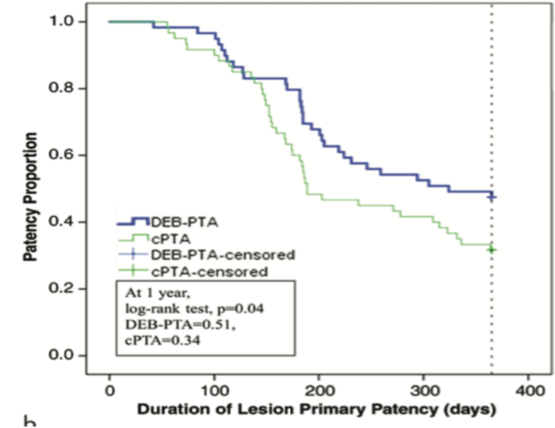

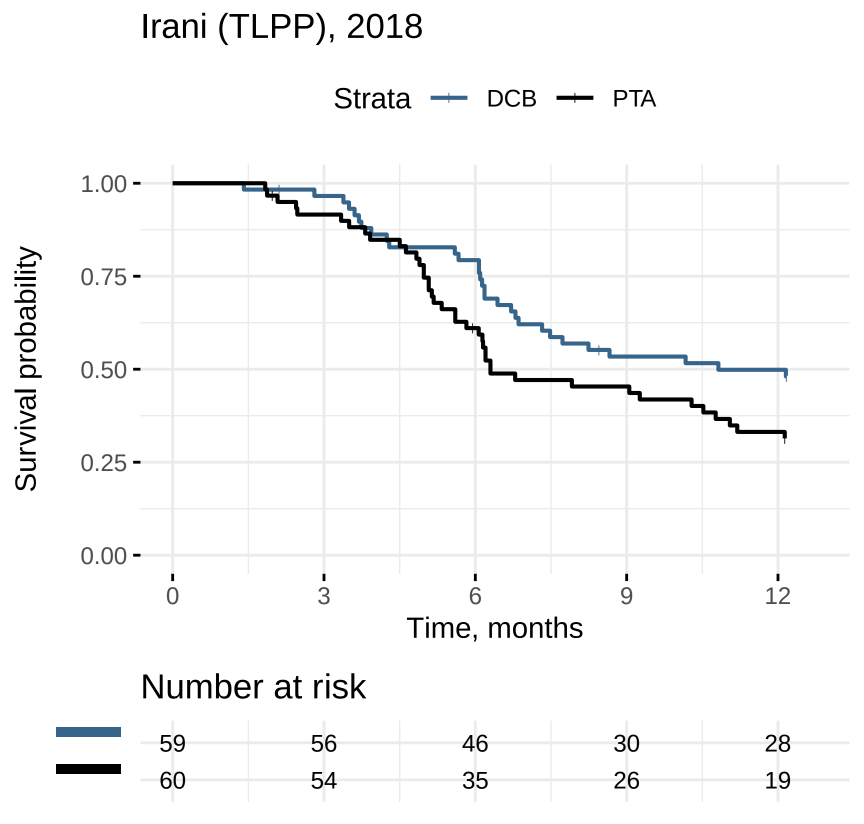
Irani (TLPP), 2018
Strata
DCB
PTA
Survival probability
1.00
0.75
0.50
0.25
0.00
0
3
6
9
12
Time, months
Number at risk
59 56 46 30 28
60 54 35 26 19

### Kim (TLPP), 2020

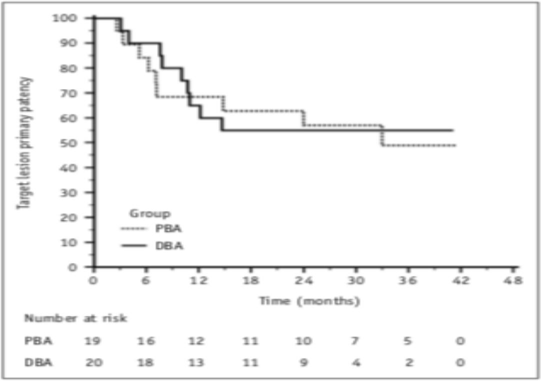


Fig. 2. Kaplan-Meier analysis of target lesion primary patency in DBA and PBA groups.

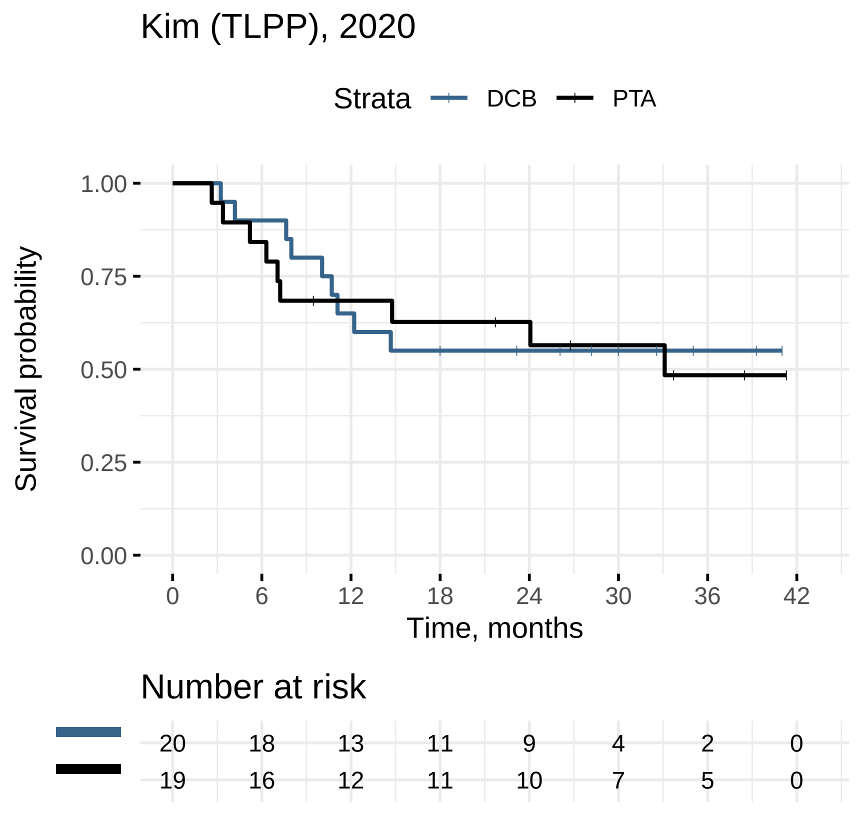

Kim (TLPP), 2020
Strata
DCB
PTA
Survival probability
1.00
0.75
0.50
0.25
0.00
0
6
12
18
24
30
36
42
Time, months
Number at risk
20 18 13 11 9 4 2 0
19 16 12 11 10 7 5 0

### Kitrou–A (TLPP), 2015

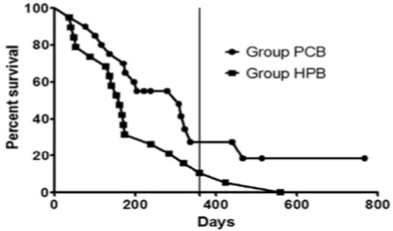


| | | | | | | |
|---|---|---|---|---|---|---|
| Group HPB | 20 | 12 | 5 | 1 | 1 | Subjects at risk |
| Group PCB | 20 | 6 | 2 | 0 | 0 | |

**Figure 2.** Kaplan–Meier survival plots of TLR-free survival. Vertical line represents 1-year time point. Subjects at risk are also presented.

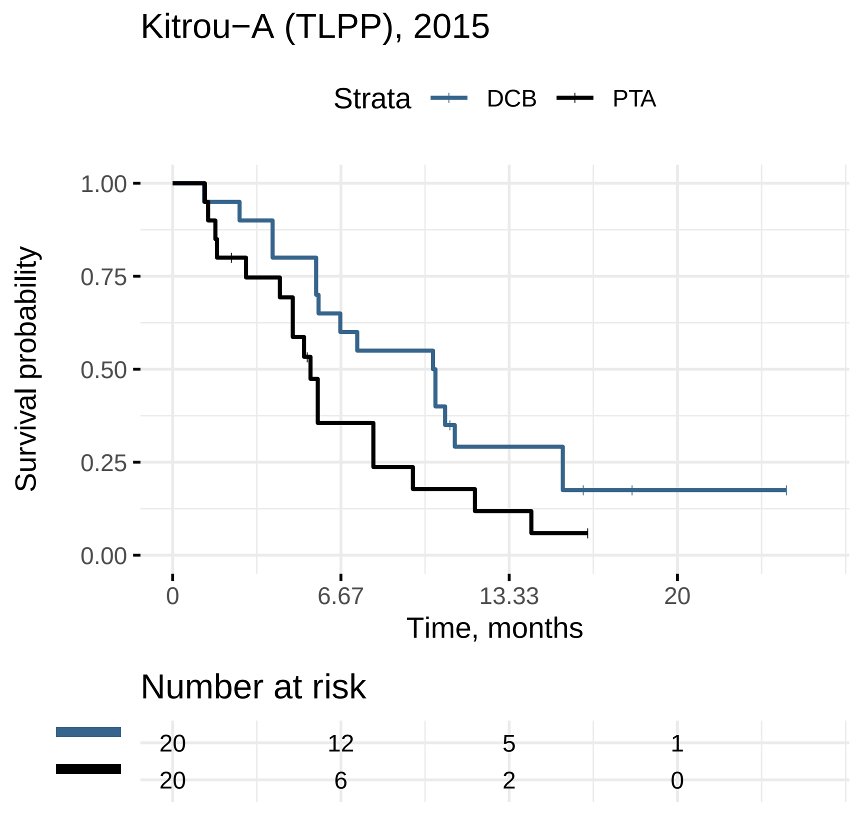
Kitrou−A (TLPP), 2015
Strata
DCB
PTA
Survival probability
1.00
0.75
0.50
0.25
0.00
0
6.67
13.33
20
Time, months
Number at risk
20
12
5
1
20
6
2
0

### Kitrou–B (TLPP), 2015

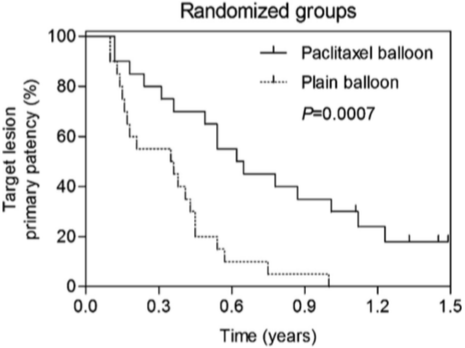


**Fig. 1.** Kaplan–Meier survival analysis of target lesion primary patency of all cases randomized between DEB and BA angioplasty treatment.

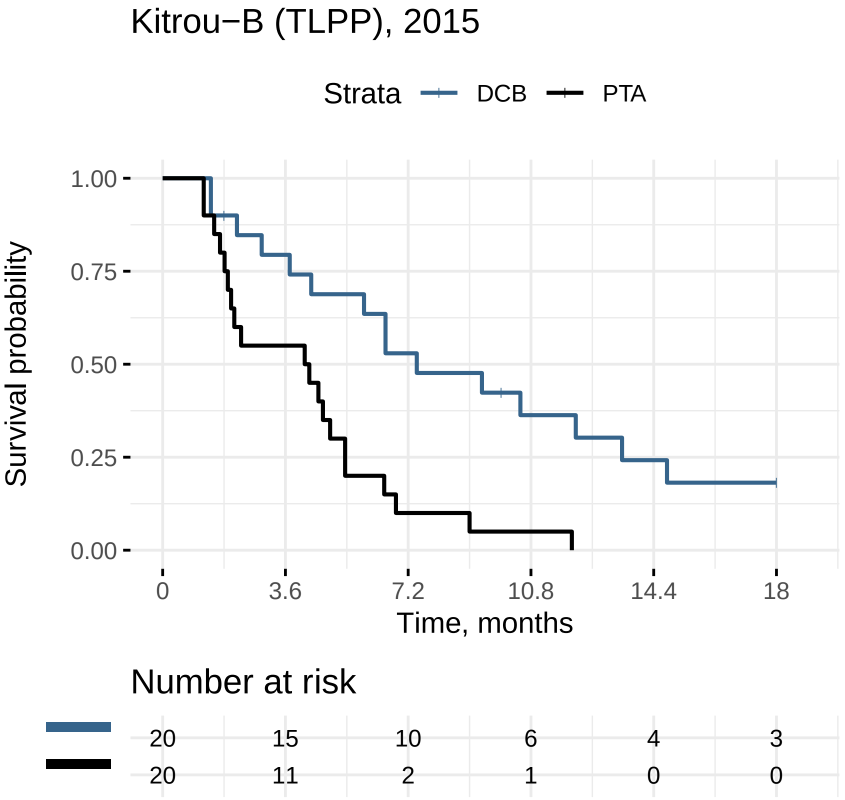

Kitrou−B (TLPP), 2015
Strata
DCB
PTA
Survival probability
1.00
0.75
0.50
0.25
0.00
0
3.6
7.2
10.8
14.4
18
Time, months
Number at risk
20 15 10 6 4 3
20 11 2 1 0 0

### Liao (TLPP), 2019

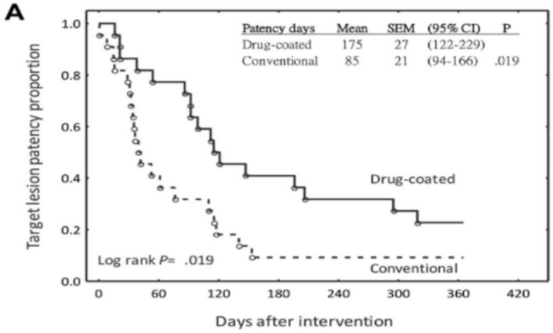


**Number at risk**

| | | | | | | | |
|---|---|---|---|---|---|---|---|
| Drug-coated | 22 | 17 | 11 | 9 | 7 | 6 | 5 |
| Conventional | 22 | 9 | 4 | 2 | 2 | 2 | 2 |

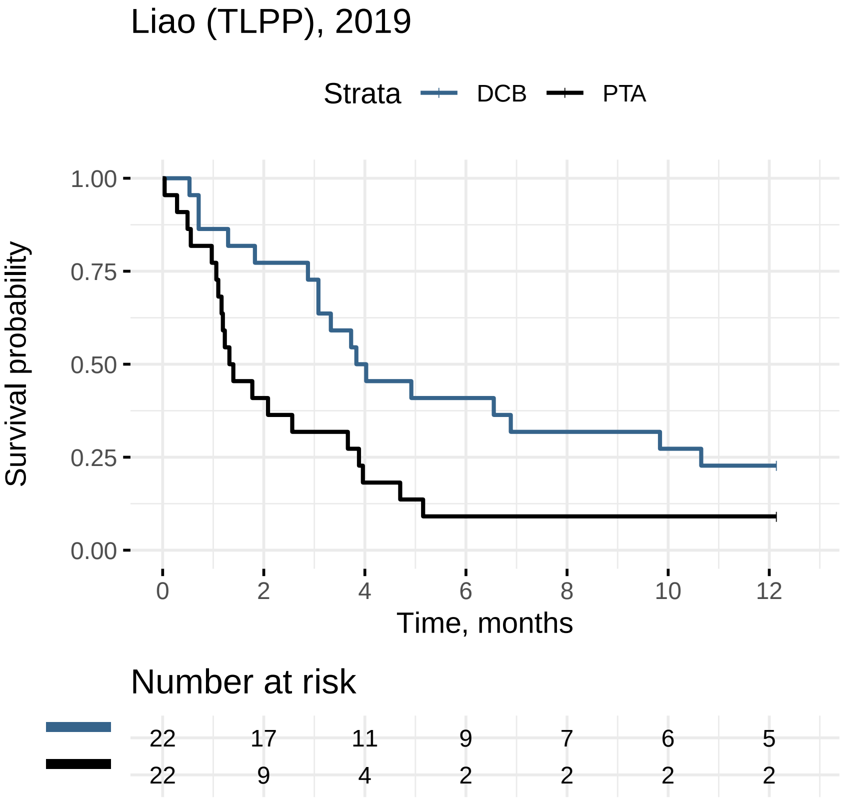
Liao (TLPP), 2019
Strata
DCB
PTA
1.00
0.75
0.50
0.25
0.00
Survival probability
0
2
4
6
8
10
12
Time, months
Number at risk
22 17 11 9 7 6 5
22 9 4 2 2 2 2

### Lookstein (TLPP), 2020

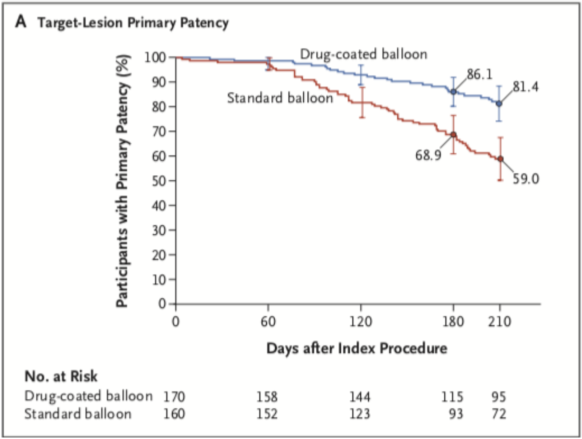

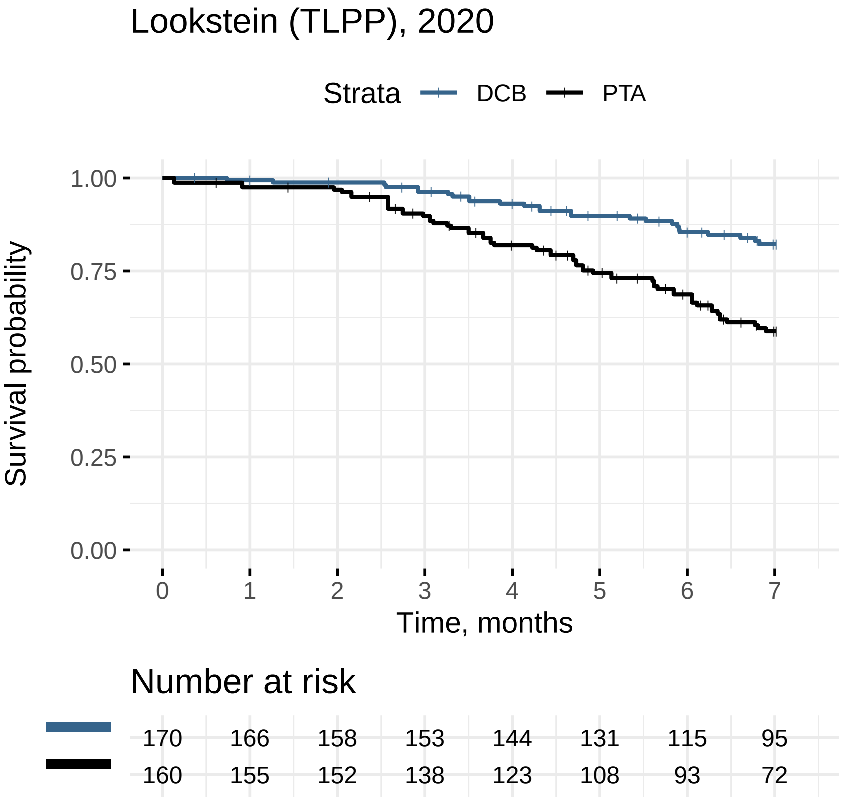
Lookstein (TLPP), 2020
Strata
DCB
PTA
Survival probability
1.00
0.75
0.50
0.25
0.00
0
1
2
3
4
5
6
7
Time, months
Number at risk
170 166 158 153 144 131 115 95
160 155 152 138 123 108 93 72

### Morenosanchez (TLPP), 2020

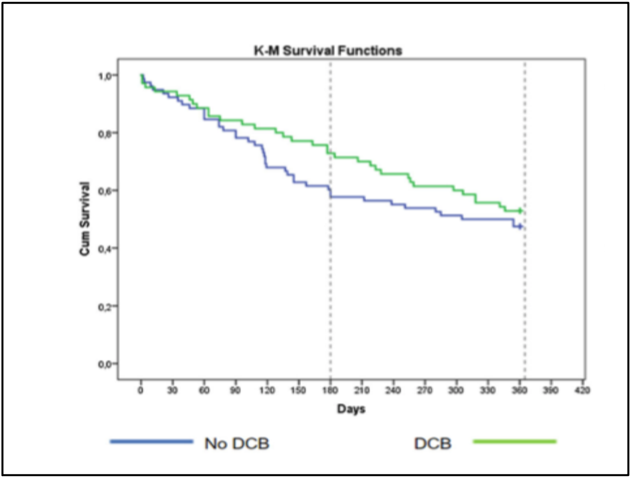

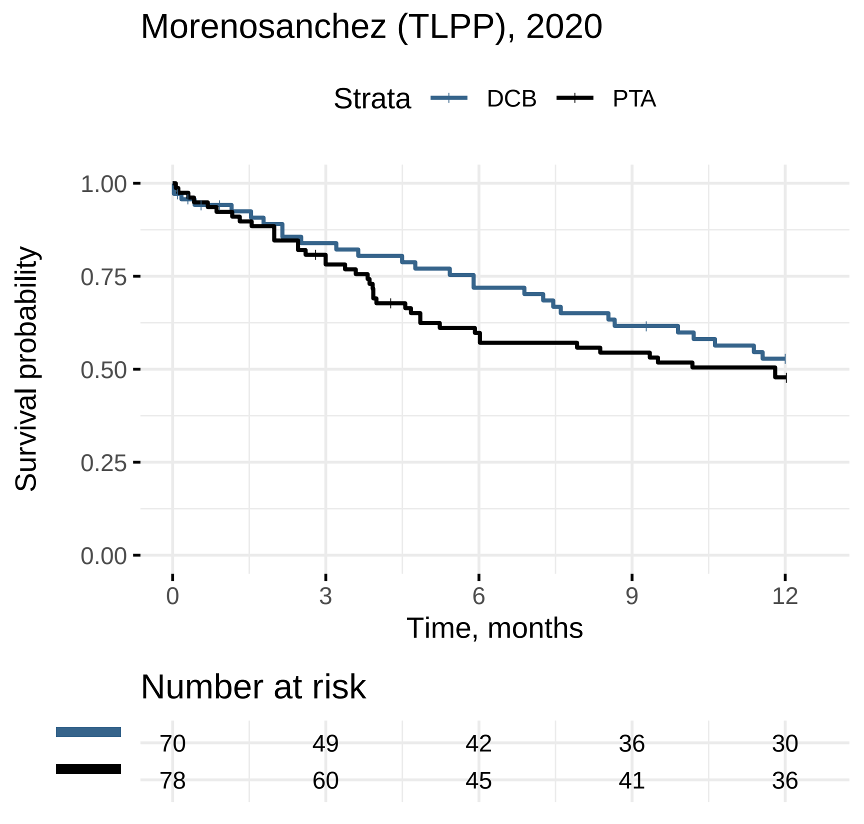
Morenosanchez (TLPP), 2020
Strata
DCB
PTA
1.00
0.75
0.50
0.25
0.00
Survival probability
0
3
6
9
12
Time, months
Number at risk
70 49 42 36 30
78 60 45 41 36

### Roosen (TLPP), 2017

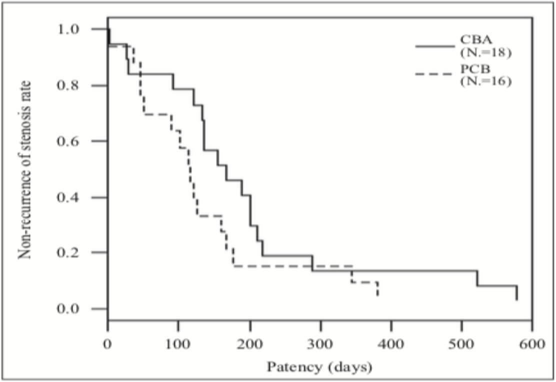


Figure 1.—Kaplan Meier Survival Curve, the patency of the PCB and CBA group.

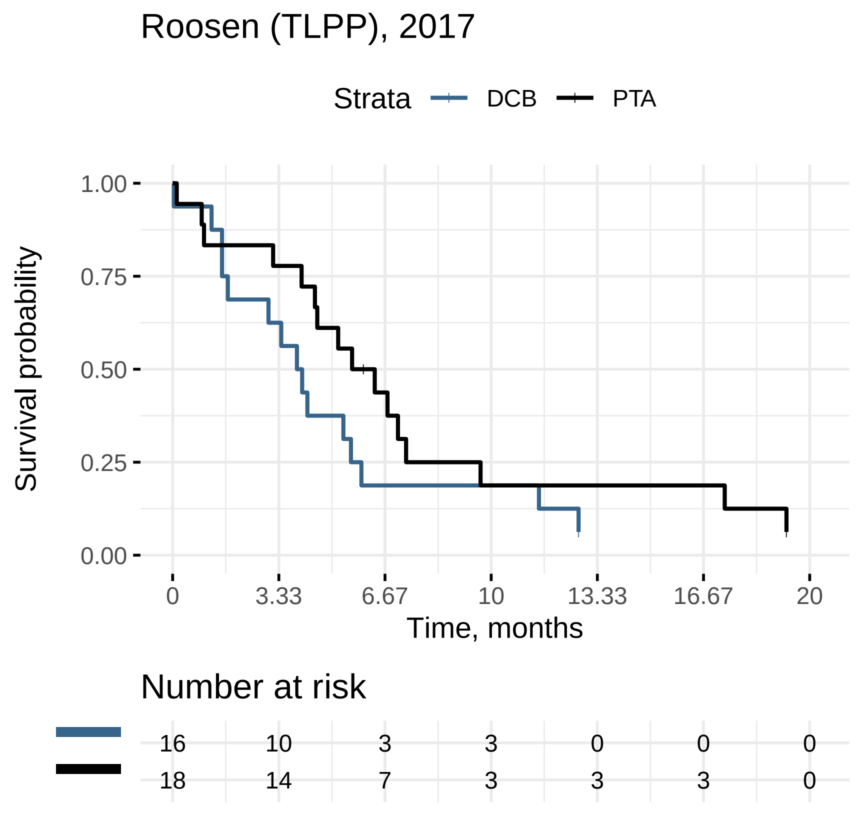

Roosen (TLPP), 2017
Strata
DCB
PTA
Survival probability
1.00
0.75
0.50
0.25
0.00
0
3.33
6.67
10
13.33
16.67
20
Time, months
Number at risk
16 10 3 3 0 0 0
18 14 7 3 3 3 0

### Swinnen (TLPP), 2018

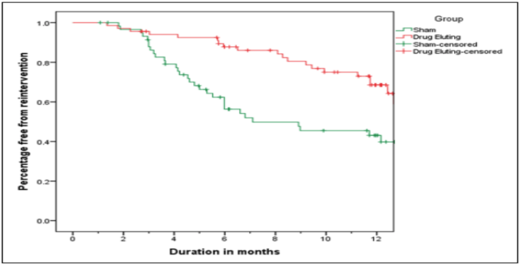


N at risk

| Group | | | | | | | |
|---|---|---|---|---|---|---|---|
| DEB | 68 | 66 | 61 | 52 | 47 | 39 | 25 |
| Sham | 60 | 56 | 44 | 28 | 23 | 20 | 14 |

**Figure 4.** Kaplan–Meier curve for freedom from reintervention to 12 months.

Swinnen (TLPP), 2018

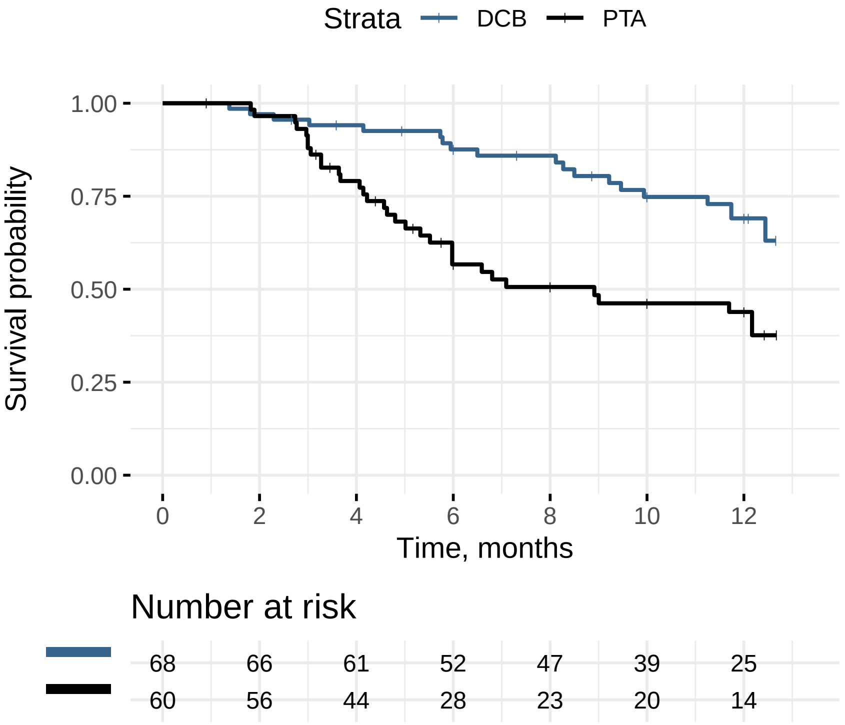

### Trerotola (TLPP), 2019

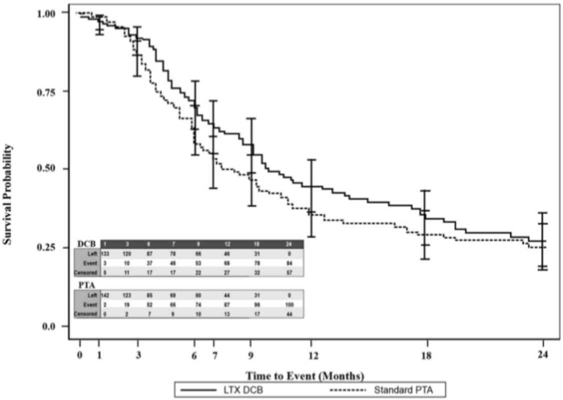


**Figure 2.** TLPP between the LTX DCB and PTA (control).

Trerotola (TLPP), 2019

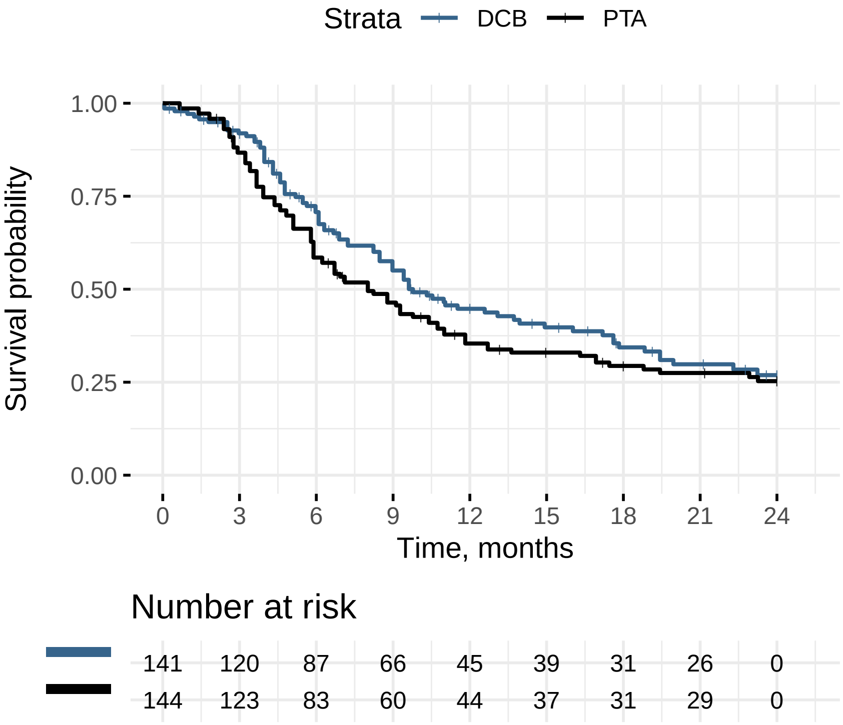
