## Supplementary Comparisons 2 for "Drug-coated balloons for dysfunctional hemodialysis venous access: A patient-level meta-analysis of randomized controlled trials"

### Bjorkman (ACPP), 2018

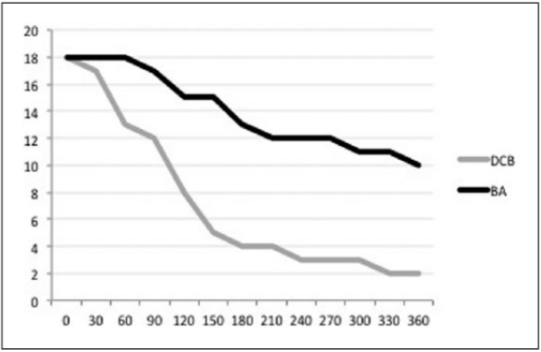


Fig. 2. Survival plot for intervention-free survival.

Bjorkman (ACPP), 2018

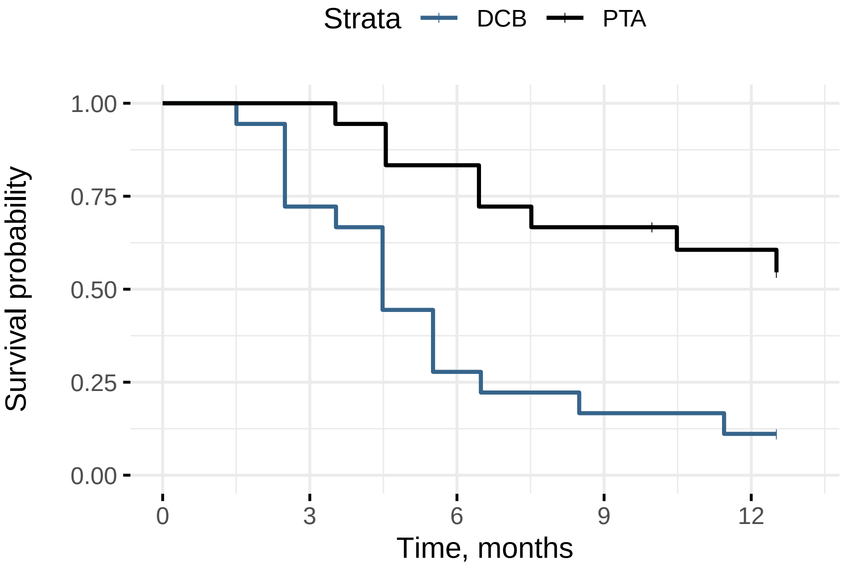


Number at risk

| | | | | |
|---|---|---|---|---|
| 18 | 13 | 5 | 3 | 2 |
| 18 | 18 | 15 | 12 | 10 |

### Irani (ACPP), 2018

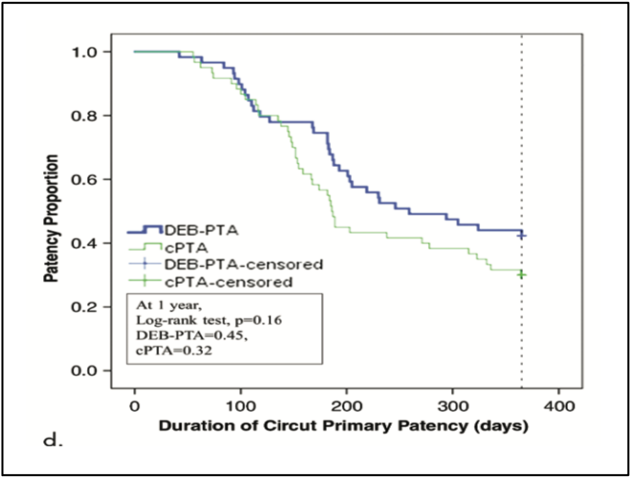

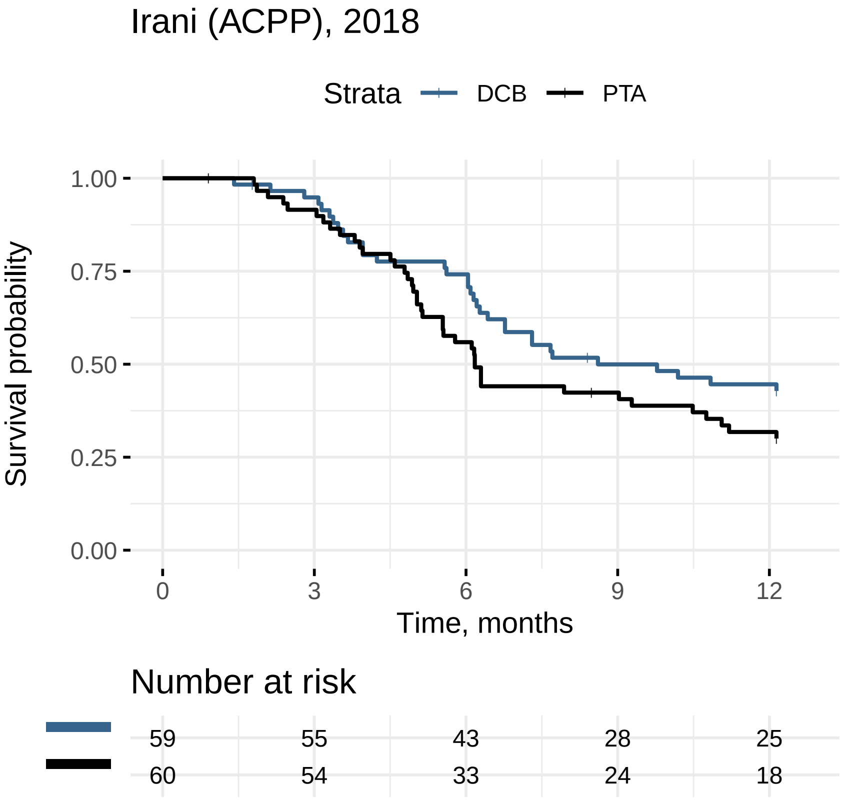
Irani (ACPP), 2018
Strata
DCB
PTA
Survival probability
1.00
0.75
0.50
0.25
0.00
0
3
6
9
12
Time, months
Number at risk
59
55
43
28
25
60
54
33
24
18

### Kitrou−A (ACPP), 2015

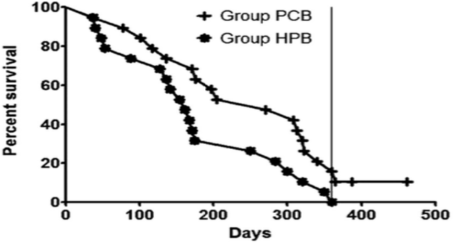


| | | | | | | |
|---|---|---|---|---|---|---|
| Group HPB | 20 | 17 | 12 | 9 | 1 | Subjects at risk |
| Group PCB | 20 | 15 | 7 | 4 | 0 | |

**Figure 3.** Kaplan–Meier survival plots of circuit primary patency. Vertical line represents 1-year time point. Subjects at risk are presented for intervals of 100, 200, 300, and 400 days.

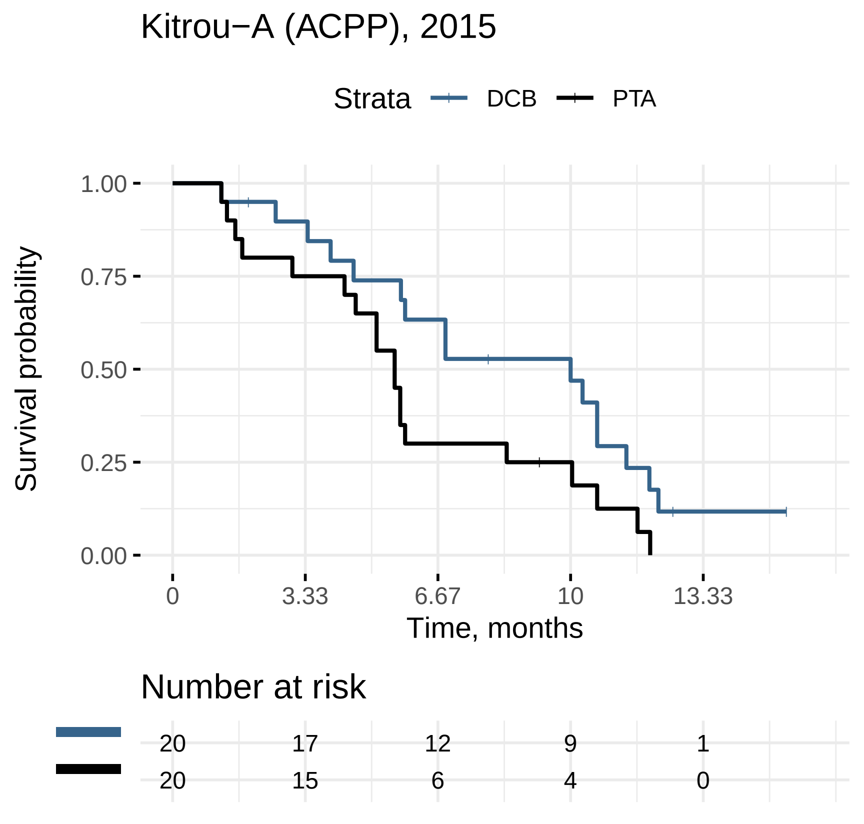
Kitrou−A (ACPP), 2015
Strata
DCB
PTA
Survival probability
1.00
0.75
0.50
0.25
0.00
0
3.33
6.67
10
13.33
Time, months
Number at risk
20 17 12 9 1
20 15 6 4 0

### Liao (ACPP), 2019

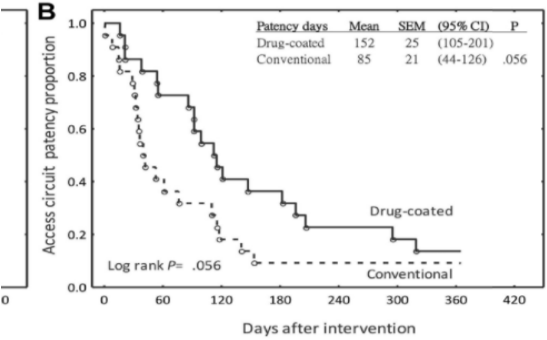


**Number at risk**

| | | | | | | | |
|---|---|---|---|---|---|---|---|
| Drug-coated | 22 | 16 | 15 | 8 | 5 | 4 | 3 |
| Conventional | 22 | 9 | 4 | 2 | 2 | 2 | 2 |

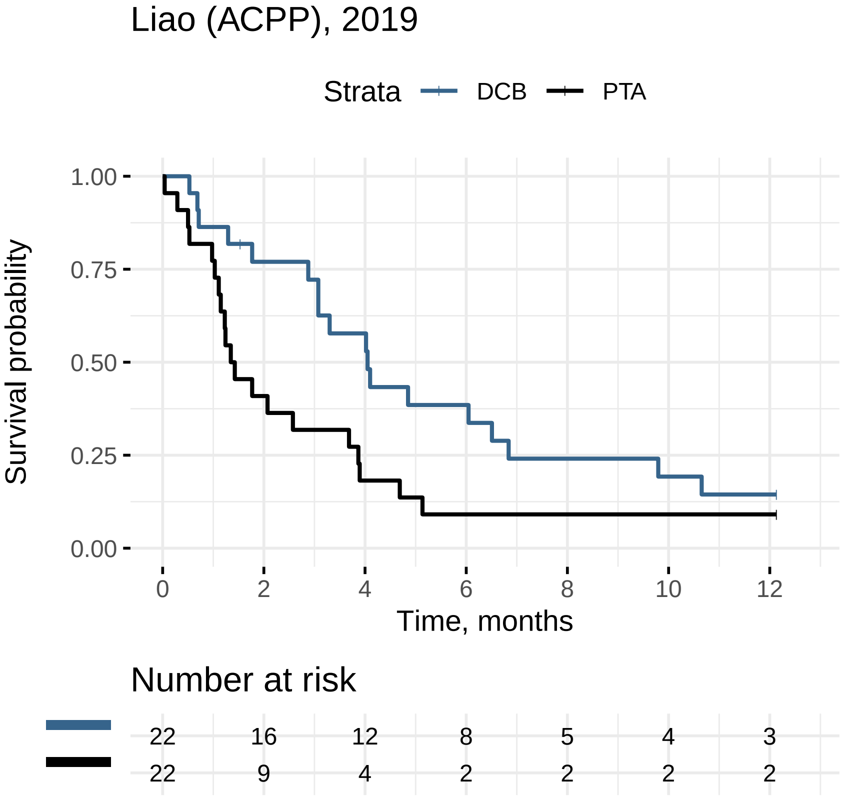

Liao (ACPP), 2019
Strata
DCB
PTA
Survival probability
1.00
0.75
0.50
0.25
0.00
0
2
4
6
8
10
12
Time, months
Number at risk
22 16 12 8 5 4 3
22 9 4 2 2 2 2

### Lookstein (ACPP), 2020

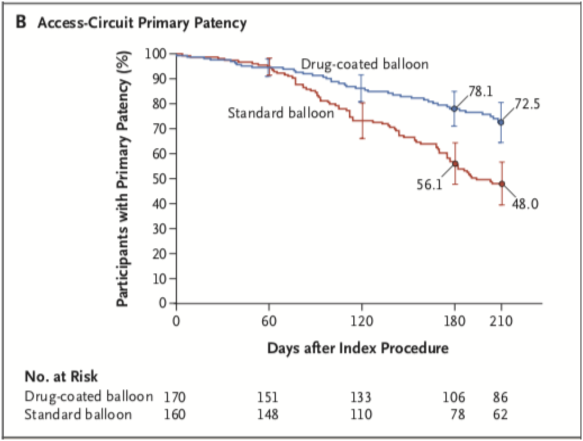

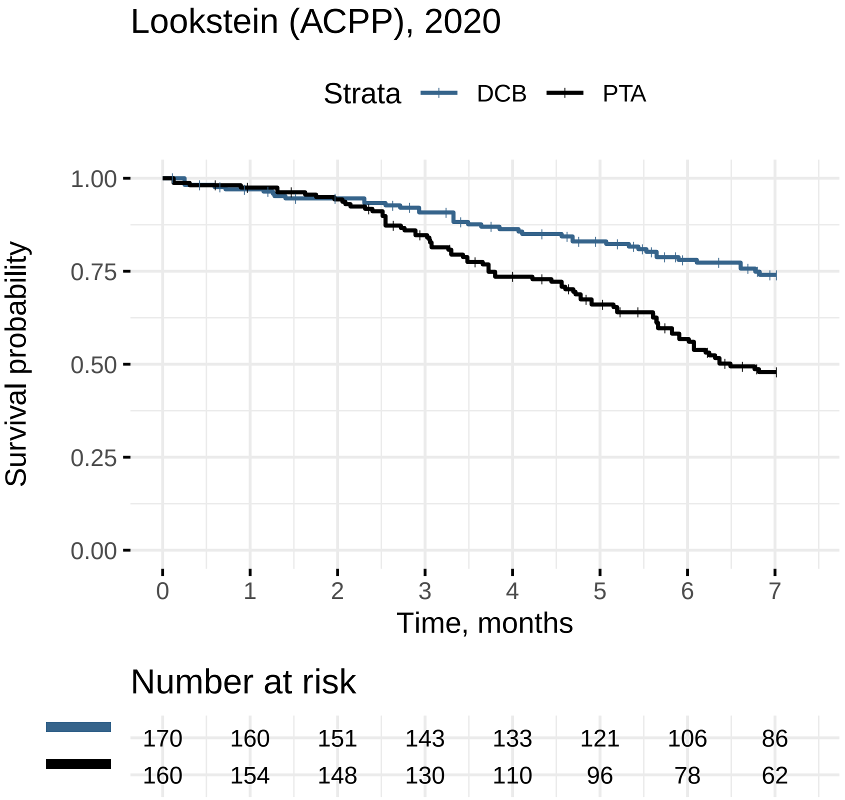

Lookstein (ACPP), 2020
Strata
DCB
PTA
Survival probability
1.00
0.75
0.50
0.25
0.00
0
1
2
3
4
5
6
7
Time, months
Number at risk
170 160 151 143 133 121 106 86
160 154 148 130 110 96 78 62

### Trerotola (ACPP), 2019

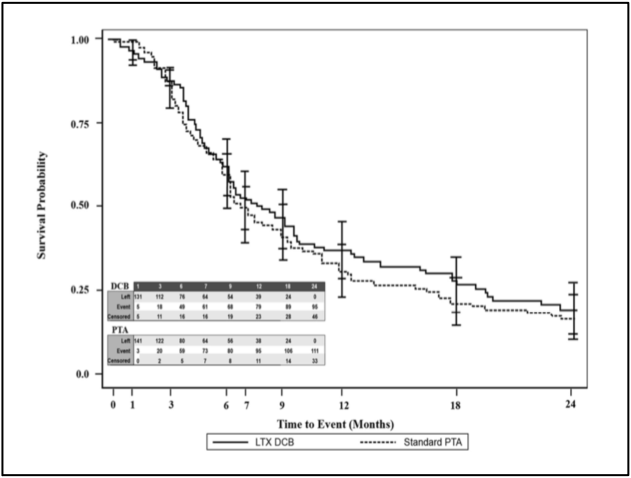

### Trerotola (ACPP), 2019

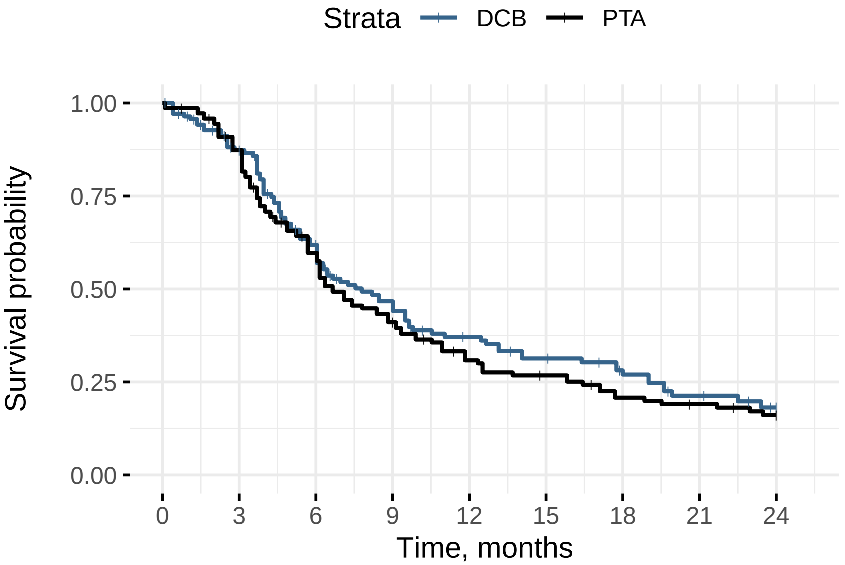


#### Number at risk

| | | | | | | | | | |
|---|---|---|---|---|---|---|---|---|---|
| | 141 | 112 | 76 | 54 | 39 | 32 | 24 | 18 | 0 |
| | 144 | 122 | 80 | 55 | 38 | 32 | 24 | 20 | 0 |
